# How Best to Explain Machine Learning Models to Clinicians: A User Study of Explanation Types

**DOI:** 10.64898/2026.06.30.26356761

**Authors:** Bowman Brown, Madeline Oguss, Kyle A. Carey, Jennifer Martin, Charles A. Kotula, Oliver T. Nguyen, Mary Akel, Douglas A. Wiegmann, Dana P. Edelson, Anoop Mayampurath, Matthew M. Churpek, Mark Craven

**Author notes:** **Corresponding Author:** Bowman Brown, Department of Computer Sciences, University of Wisconsin-Madison.

## Abstract

**Background:** Explanations play a crucial role in helping clinicians understand how black-box machine learning models make predictions in clinical settings. Several different types of explanations have been developed, each corresponding to a unique approach for characterizing the relationships between model inputs and predictions. However, it remains unclear what types of explanations are the most valued by clinicians.

**Objective:** To improve the utility of machine learning in clinical settings, we aimed to evaluate how different explanation methods are valued by clinicians across clinically important metrics, such as importance, trust, understanding, and how explanations affect clinicians’ thinking about patients.

**Methods:** We conducted a user study of 39 critical care and hospital medicine nurses and physicians to compare attribution, counterfactual, and rule-based explanations. We analyzed the impact of each type of explanation on clinicians’ trust in and understanding of the predictions made by machine learning models, how well clinicians understood the explanation, and how the explanation affected what they thought were the most important features for determining patients’ status. We also assessed clinicians’ preferences for the representation of different types of explanations.

**Results:** Clinicians consider explanations of clinical machine learning models important, with physicians perceiving explanations as more important after interacting with them than nurses. All explanation types affected clinicians across all measured dimensions, with attribution explanations having the most significant positive effects on all measured dimensions. Moreover, nearly half of clinicians preferred viewing multiple explanation types together.

**Conclusions:** It is important to provide explanations for predictions made by machine learning models in clinical settings. When implementing machine learning explanations in these settings, developers should prioritize attribution explanations while allowing for multiple types of explanations to be shown. Furthermore, the development of new explanation methods should be tailored towards specific clinical roles, as nurses and physicians may utilize explanations differently to support their respective workflows.

## Introduction

Machine learning (ML) models are increasingly used in clinical settings, with a survey by the American Medical Association finding that in late 2024, approximately 66% of physicians used some form of ML in clinical practice [1]. Although clinicians report a desire to understand how ML models make predictions [2,3], many types of ML models used in clinical practice, such as neural networks, gradient-boosted trees, and random forests, are challenging for a clinician to interpret [4].

Explainable Artificial Intelligence (XAI) methods convey information about how ML models make predictions through *explanations*. Explanations are simplified representations of a model’s prediction process that relate how input feature values affect the output predictions [5]. For example, an explanation might provide a numeric value for each feature, indicating how much that feature contributed to the output, or may suggest that if the value for a feature at a particular time point were to change, the output would also change.

Effective use of explanations in clinical settings requires understanding how different explanations are valued by clinicians. Although explanations of ML models have been shown to affect clinician decision-making [6,7], there remains a gap in understanding which specific *explanation types* are most valued by clinicians. Each explanation type corresponds to a unique approach for characterizing the relationship between input feature values and a model’s predictions, potentially affecting clinicians’ thinking in different ways. Determining the explanation types that are most valued by clinicians can lead to changes in ML-assisted clinical practice and improve patient outcomes.

To address this gap, we conducted a user study of nurses and physicians to compare three of the most prominent explanation types: *attribution*, *counterfactual*, and *rule-based*. Figure 1 provides an example of each explanation type. An attribution explanation indicates how much each part of the input instance contributes to the prediction for that instance. A counterfactual explanation shows how to modify the input instance to change the prediction for that instance. A rule-based explanation identifies the conditions satisfied by the input instance that result in the prediction for that instance. We focused on these explanation types as they are widely used and provide users with information regarding the model’s prediction process.

**Figure 1.**
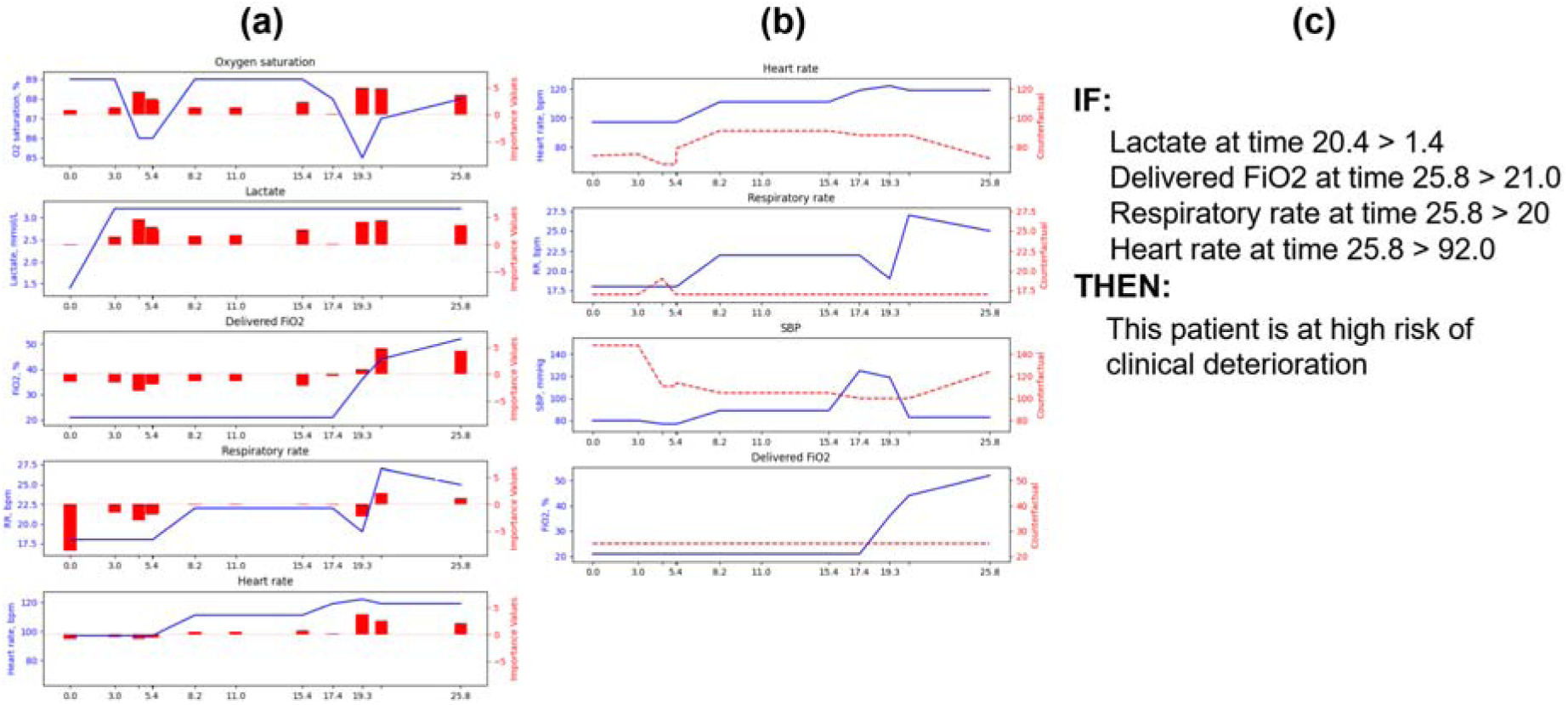
Examples of the different explanation types in our user study. (a) An attribution explanation: each line plot represents a distinct feature, where the blue line denotes the feature measurements over time and the red bars indicate the importance values computed by the attribution explanation for each feature at each time point. (b) A counterfactual explanation: each line plot represents a distinct feature, where the blue line denotes the feature measurements over time and the red dotted line indicates the modified feature values that would change the prediction. (c) A rule-based explanation: a list of conditions specifying tests on features is shown under the IF header, with the prediction for which the conditions are sufficient listed under the THEN header.

All of the explanation types compared in this work are *local*, *post-hoc*, *model-agnostic* explanations, corresponding to the most broadly applicable types of explanations and covering common approaches to explaining ML models in clinical settings. A local explanation provides information about how the ML model makes a prediction for a single input instance, such as for a specific patient. A post-hoc approach explains a model after the model has been fit to data. A model-agnostic approach can be applied to models of any type [5,8]. Figure 2 shows the process of generating local, post-hoc, model-agnostic explanations.

**Figure 2:**
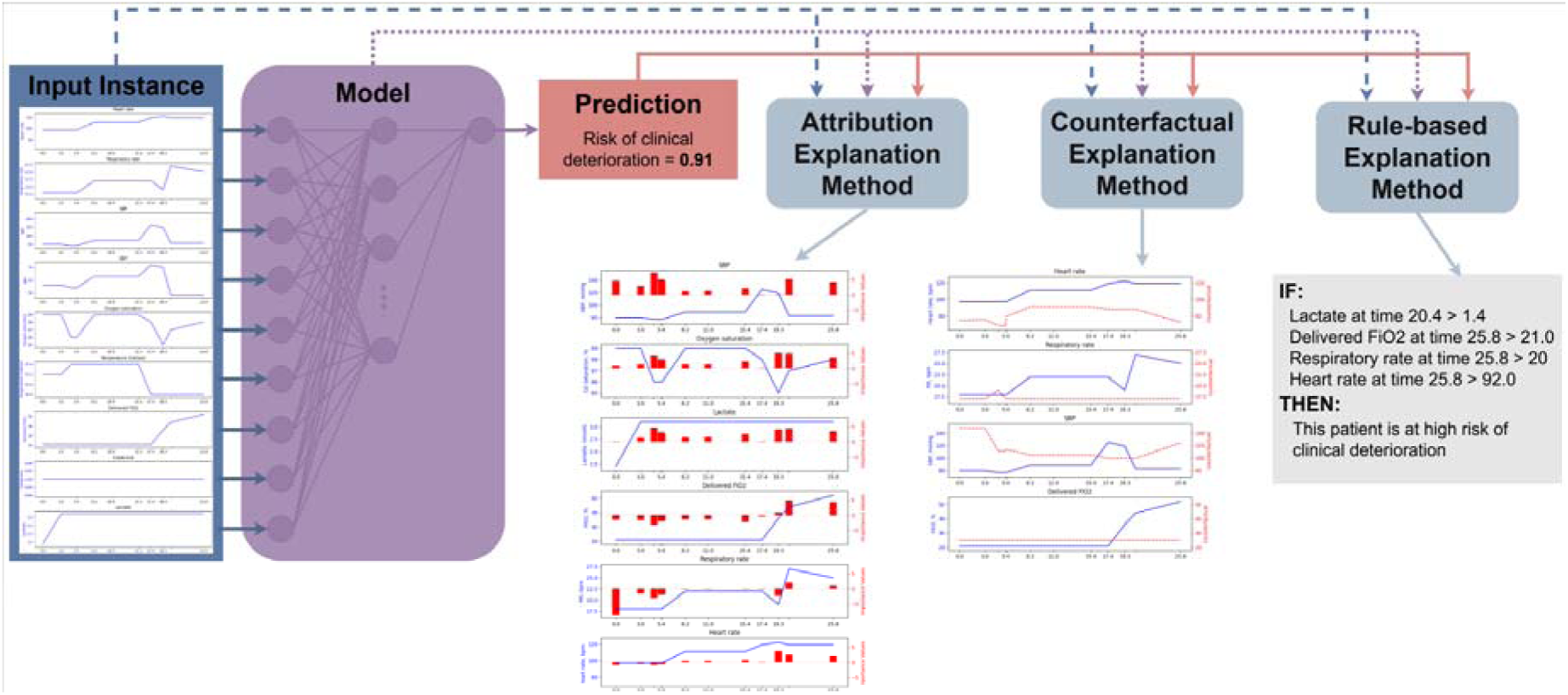
The process of generating a local post-hoc explanation. An input instance is passed into a model to make a prediction. The input instance, model, and prediction are all used by each explanation method to generate an explanation of a given type.

To understand the value of different types of explanations to clinicians, we assess the importance that clinicians assign to explanations in general (*explanation importance*) and evaluate the effects of attribution, counterfactual, and rule-based explanation types on four key dimensions: (1) trust in the predictions of the ML model (*model trust*), (2) understanding of how the ML model is making the predictions (*model understanding*), (3) understanding of a given explanation (*explanation understanding*), and (4) understanding of the features important for determining a patient’s status (*influence*). In addition, we measure clinicians’ preferences for the representation of different explanation types.

Our user study is unique in that we compare attribution, counterfactual, and rule-based explanation types consistent with their use in real clinical settings: We apply the XAI methods Integrated Gradients [9] to generate attribution explanations, CoMTE [10] to generate counterfactual explanations, and Anchors [11] to generate rule-based explanations for a deep learning model trained on longitudinal (time series) patient data to predict risk of clinical deterioration. We had nurses and physicians assess explanations of all three types for five synthetic patient cases and answer questions intended to measure explanation importance, model trust, model understanding, explanation understanding, influence, and their preferences for explanation representation.

The contributions of this paper are:

1. An in-depth evaluation of explanations generated using XAI methods for the predictions of a real clinical deep learning model across multiple essential dimensions for determining the value of explanations in clinical settings.
2. A comparison of different explanation types to identify how each type of explanation affects each measured dimension, and where certain explanation types might be of most value to clinicians.
3. Guidance based on clinician feedback for implementing explanations in clinical settings.

Our results reveal that explanations in the context of early identification of clinical deterioration are considered important by clinicians, and that all three explanation types increase clinicians’ model trust, increase model understanding, are understood by clinicians, and influence clinicians’ thinking about patient cases. In particular, attribution explanations show the most significant effects across all four dimensions compared to counterfactual and rule-based explanations. Furthermore, clinicians generally prefer to see multiple explanation types together and prefer more concise representations.

### Related Work

We define related work as user studies that evaluate at least one type of explanation. We divide these studies into four categories based on whether they compare multiple explanation types or not, and whether they focus on clinical tasks or not.

Many user studies of explanation methods do not focus on comparing multiple explanation types or on clinical domains. The survey by Rong et al. [12] provides a good overview of such studies.

A number of user studies aim to determine how different explanation types affect general users. Our study differs from these works in that it focuses on a clinical domain and uses clinicians as study participants. None of the identified works compares all three of attribution, counterfactual, and rule-based explanation types or measures all of the key dimensions of model trust, model understanding, explanation understanding, and influence [13–22]. Additionally, only four of these works [13,17,18,21] use actual ML models to make predictions and XAI methods to generate explanations, and of these, only Jeyakumar et al. [17] and Chae et al. [21] use models trained on data where instances include measurements from more than one time point.

Several user studies recruit clinicians, but do not compare different explanation types [23–28]. These user studies evaluate a single explanation type and do not measure at least one of the dimensions of model trust, model understanding, explanation understanding, or influence.

The user studies that most closely align with our work compare different explanation types in user studies of clinicians [29–32]. However, none of the related works use data where instances include measurements from more than one time point, and all of them differ in at least one other meaningful way. Jacobs et al. [29] compare rule-based and attribution explanations, measuring clinicians’ self-reported influence for each explanation type. However, they do not consider counterfactual explanations, do not explain the predictions of an actual model, and do not assess model trust, model understanding, or explanation understanding. Bergomi et al. [30] compare rule-based and attribution explanations, measure the influence of each explanation type, and use ML models and XAI methods for their user study rather than manually constructing predictions or explanations. However, they do not measure clinicians’ model trust or model understanding. Hou et al. [31] omit counterfactual and rule-based methods from their comparison and do not report the model understanding or influence of each explanation type. While they use a similar ICU clinical deterioration prediction task, they differ from our work by using a model trained on clinical notes instead of tabular data.

The work most similar to ours is that of Schoonderwoerd et al. [32], who compare the effects of attribution, counterfactual, and rule-based explanations on clinicians’ model trust, model understanding, and explanation understanding. However, this study differs from ours by using manually constructed predictions and explanations and measuring only self-reported rather than actual influence on clinical tasks.

In contrast to the related work identified above, our user study is the only one to compare attribution, counterfactual, and rule-based explanation types generated by XAI methods for a real ML model across the four key dimensions with a cohort of clinicians. Additionally, our cohort of 39 nurses and physicians represents one of the largest user studies of explanations involving clinicians. Only Jacobs et al. recruited a larger total number of participants, but reported participants broadly as ‘clinicians’ without differentiating between nurses, physicians, or other types of clinicians. Our work provides the strongest evidence for understanding how all three types of explanations affect the four measured dimensions and helps identify the types of explanations with the greatest value in clinical settings.

## Methods

In this section, we detail information on the recruitment process and participants involved in the user study, the model and task for which explanations were generated, the three types of explanations, the design of the user study, and our approach for analyzing the results.

### Ethical Considerations

The study was approved by the Institutional Review Boards (IRBs) of the University of Chicago (IRB23-1734) and University of Wisconsin-Madison (#2023-1220). Each IRB waived study-specific informed consent. Study data was collected and managed using the REDCap electronic data capture tools hosted at the University of Wisconsin-Madison, School of Medicine and Public Health [33,34].

### Participant Recruitment

We recruited nurses, physician trainees, and attending physicians in critical care and hospital medicine from the University of Wisconsin-Madison and the University of Chicago Medicine health systems using email invitations. Participants received a $50 gift card incentive upon completion of the survey. In total, 62 participants started the survey, and 39 submitted completed surveys. Participants took on average 48 minutes to complete the survey. We filtered out incomplete surveys and used only the 39 completed surveys in our analysis. Details about the participant demographics are shown in Table 1.

**Table 1.**
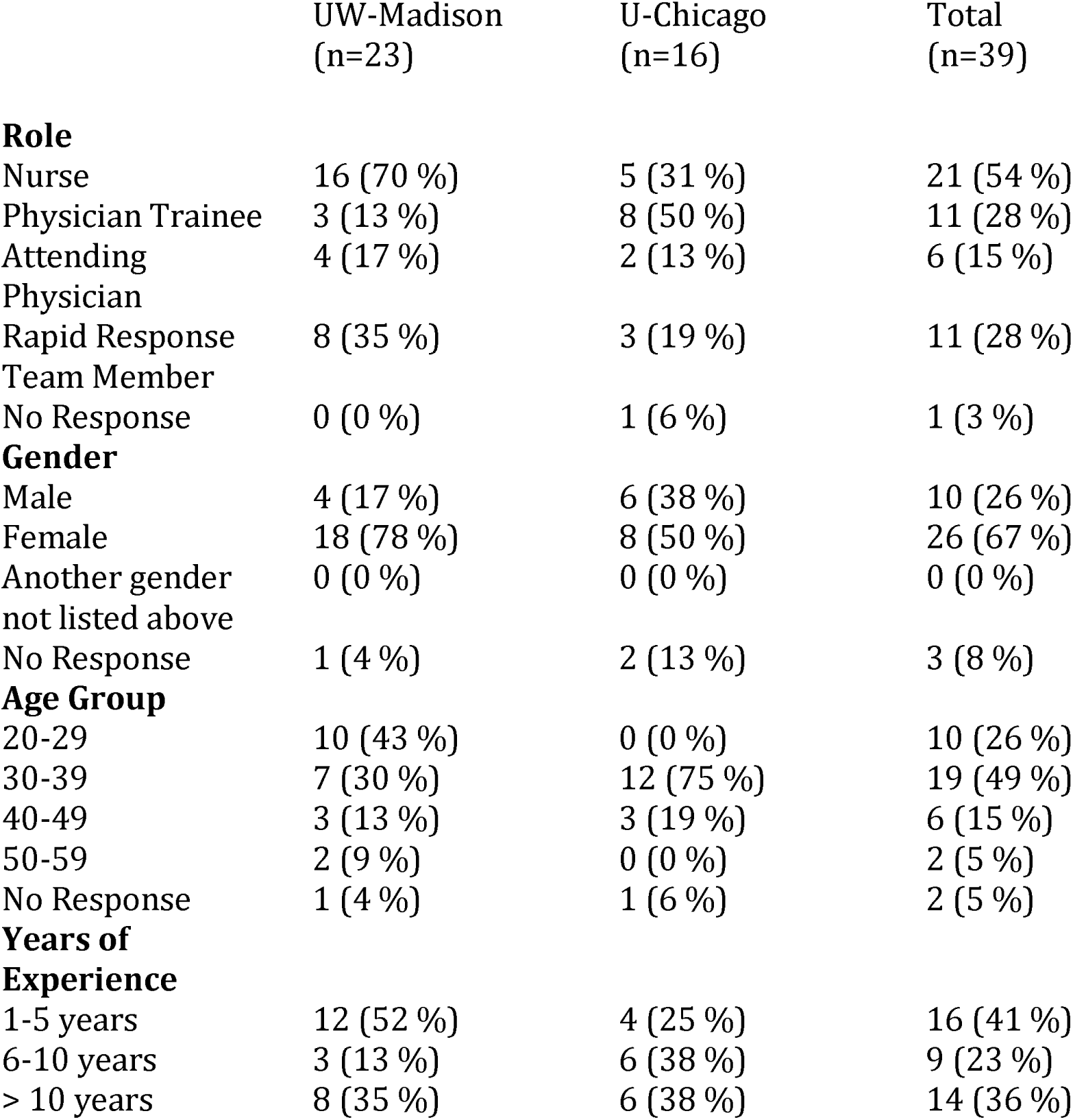
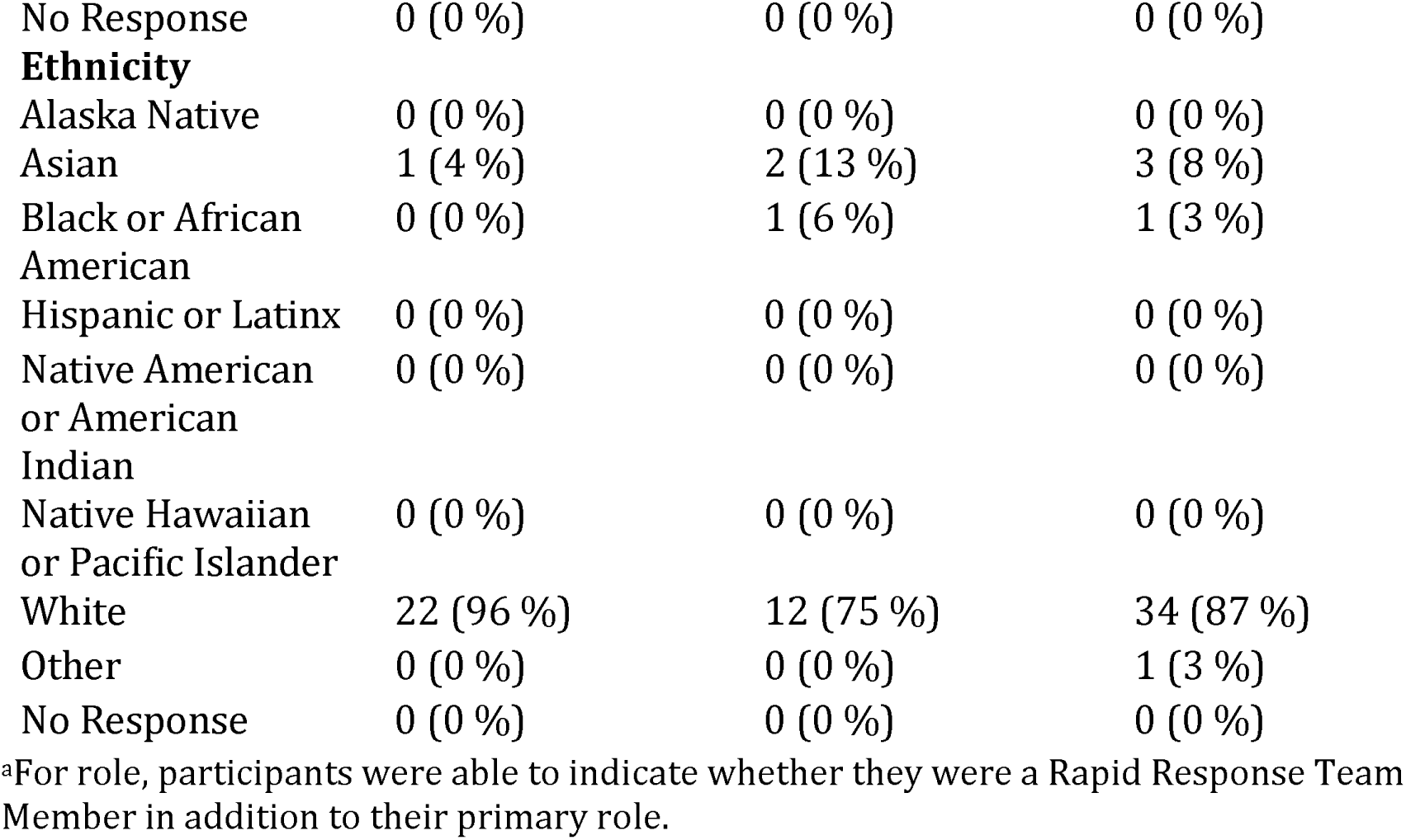
User study participant demographics by site.^a^.

|  | UW-Madison<br>(n=23) | U-Chicago<br>(n=16) | Total<br>(n=39) |
| --- | --- | --- | --- |
| **Role** |  |  |  |
| Nurse | 16 (70 %) | 5 (31 %) | 21 (54 %) |
| Physician Trainee | 3 (13 %) | 8 (50 %) | 11 (28 %) |
| Attending Physician | 4 (17 %) | 2 (13 %) | 6 (15 %) |
| Rapid Response Team Member | 8 (35 %) | 3 (19 %) | 11 (28 %) |
| No Response | 0 (0 %) | 1 (6 %) | 1 (3 %) |
| **Gender** |  |  |  |
| Male | 4 (17 %) | 6 (38 %) | 10 (26 %) |
| Female | 18 (78 %) | 8 (50 %) | 26 (67 %) |
| Another gender not listed above | 0 (0 %) | 0 (0 %) | 0 (0 %) |
| No Response | 1 (4 %) | 2 (13 %) | 3 (8 %) |
| **Age Group** |  |  |  |
| 20-29 | 10 (43 %) | 0 (0 %) | 10 (26 %) |
| 30-39 | 7 (30 %) | 12 (75 %) | 19 (49 %) |
| 40-49 | 3 (13 %) | 3 (19 %) | 6 (15 %) |
| 50-59 | 2 (9 %) | 0 (0 %) | 2 (5 %) |
| No Response | 1 (4 %) | 1 (6 %) | 2 (5 %) |
| **Years of Experience** |  |  |  |
| 1-5 years | 12 (52 %) | 4 (25 %) | 16 (41 %) |
| 6-10 years | 3 (13 %) | 6 (38 %) | 9 (23 %) |
| > 10 years | 8 (35 %) | 6 (38 %) | 14 (36 %) |
| No Response | 0 (0 %) | 0 (0 %) | 0 (0 %) |
| **Ethnicity** |  |  |  |
| Alaska Native | 0 (0 %) | 0 (0 %) | 0 (0 %) |
| Asian | 1 (4 %) | 2 (13 %) | 3 (8 %) |
| Black or African American | 0 (0 %) | 1 (6 %) | 1 (3 %) |
| Hispanic or Latinx | 0 (0 %) | 0 (0 %) | 0 (0 %) |
| Native American or American Indian | 0 (0 %) | 0 (0 %) | 0 (0 %) |
| Native Hawaiian or Pacific Islander | 0 (0 %) | 0 (0 %) | 0 (0 %) |
| White | 22 (96 %) | 12 (75 %) | 34 (87 %) |
| Other | 0 (0 %) | 0 (0 %) | 1 (3 %) |
| No Response | 0 (0 %) | 0 (0 %) | 0 (0 %) |

<sup>a</sup>For role, participants were able to indicate whether they were a Rapid Response Team Member in addition to their primary role.

### Model and Task

We generated explanations for a set of predictions made by a model similar to eCARTv5 [35], which has previously been implemented in clinical practice. Our model outputs a clinical deterioration risk score corresponding to a patient’s likelihood of death or ICU transfer from the medical-surgical wards within 24 hours of the prediction time. In contrast to eCARTv5, our model is based on the LSTM [36] deep neural network architecture.

Model training followed the process given by Kotula et al. [37]. The model was trained on 284,302 adult inpatient admissions with ward stays collected from 2016 to 2022 at the University of Chicago. Input data for the model consists of a patient trajectory represented by 55 demographic, laboratory measurement, and vital sign structured features recorded at various time points after admission.

We used five synthetic patient cases for the user study. First, we selected a random subset of five de-identified patient cases in the 95th risk percentile from the model’s training data. For each case, we perturbed a random subset of 10% of time points in 10% of features by adding or subtracting uniformly sampled noise to increase deterioration risk, depending on whether higher or lower values increased risk. Perturbations were limited so as not to exceed the maximum or minimum values observed in the training data for each feature. The five synthetic cases were validated by an expert clinician (M.M.C.) to ensure they were realistic and perceived to be high risk.

### Explanation Types

We generated explanations of the model’s predictions on the synthetic patient cases using three methods: Integrated Gradients [9] for attributions, CoMTE [10] for counterfactuals, and Anchors [11] for rule-based explanations. These three methods were chosen based on their recency and popularity in the research literature. A description of each explanation type and details of the specific method used to generate the explanations are given below. Examples of each explanation type can be found in Figure 1.

Given a model *f* and an input instance ***X***, we assume without loss of generality that ***X*** is a matrix of size ℝ*^K^*^×*T*^where *K* is the number of features and *T* is the number of time points. In practice, ***X*** may be a tensor of any shape. We denote the values of the *k*th feature with ***X****_k_* ∈ ℝ*^T^* and the value of the *k*th feature at the *t*th time point with *X_k_*_,*t*_ ∈ ℝ.

An attribution explanation outputs a single scalar ‘importance’ score for each *X_k_*_,*t*_. Integrated Gradients assigns an importance score to each *X_k_*_,*t*_ that represents how much the value of *X_k_*_,*t*_ changes the model’s prediction, relative to the value in some baseline instance *B_k_*_,*t*_. We define the baseline instance ***B*** to be the median of a random sample of 100 training instances. We Z-normalize the importance scores for an instance when showing them in the user study. For the example in Figure 1a, the Z-normalized Integrated Gradients explanation indicates that the value of the Delivered FiO2 feature at 25.8 hours since admission increases the predicted risk of clinical deterioration for the patient by 4.5 standard deviations more than the average risk increase given by all other features at all other time points. For Integrated Gradients explanations in the user study, we show the five features with the greatest attribution value magnitude in addition to any other features used in the counterfactual or rule-based explanations. The Integrated Gradients explanations show between 5 and 7 features (5.6 ± 0.9).

A counterfactual explanation outputs a modified version *X*^′^ of the original input instance ***X*** such that the model’s prediction for *X*^′^ crosses an implementation-defined threshold *τ*. CoMTE searches for a modified instance with the minimum number of features replaced with the values of features taken from instances in a subset of data such that the prediction for the modified instance crosses *τ*. We define *τ* as the 95th percentile of risk and the subset of data as a random sample of 100 training instances.

For the example in Figure 1b, the CoMTE explanation indicates that replacing the original instance’s Heart Rate, Respiratory Rate, SBP, and Delivered FiO2 (denoted by blue lines) with a lower Heart Rate, lower Respiratory Rate, lower Delivered FiO2 amount, and higher SBP (denoted by red dotted lines) would cause the model prediction to change from the patient being at or above the 95th percentile of risk to the patient being below the 95th percentile of risk. For CoMTE explanations in the user study, we show only the subset of modified features and their original instance values as the explanation. The CoMTE explanations show between 3 and 5 features (3.8 ± 0.8).

A rule-based explanation outputs a rule composed of a set of *n* conditions *C*_1_(***X***), …, *C_n_*(***X***) that are sufficient for the model’s prediction. Each condition *C_i_*(***X***) consists of a feature at a time point *X_k_*_,*t*_, a relational operator, and a value, and is true if the relation holds between *X_k_*_,*t*_ and the value. A rule specifies that for an input instance ***X***, for any perturbed version ***X***^′^ for which the conditions are satisfied, the model’s prediction for ***X***^′^ will not cross an implementation-defined threshold value *τ*. We define the threshold value *τ* for Anchors to be the 95th percentile of risk. For the example in Figure 1c, the Anchors explanation indicates that if Lactate was greater than 1.4 at 20.4 hours since admission and Delivered FiO2 was greater than 21, Respiratory rate was greater than 20, and Heart rate was greater than 92 at 25.8 hours since admission, changes to any other feature-time point combinations are unlikely to change the prediction from the patient being at or above the 95th percentile of risk to the patient being below the 95th percentile of risk. For Anchors explanations in the user study, rules are displayed in text as *IF [Conditions] THEN [Prediction]*. The Anchors rules show between 2 and 6 conditions (3.8 ± 1.6).

### Survey Design

Figure 3 shows an overview of the user study. Each participant completed a Pre-Cases questionnaire, a Cases questionnaire, and a Post-Cases questionnaire. All questions in the user study were optional. A copy of the questions asked in the user study can be found in the supplementary materials.

**Figure 3.**
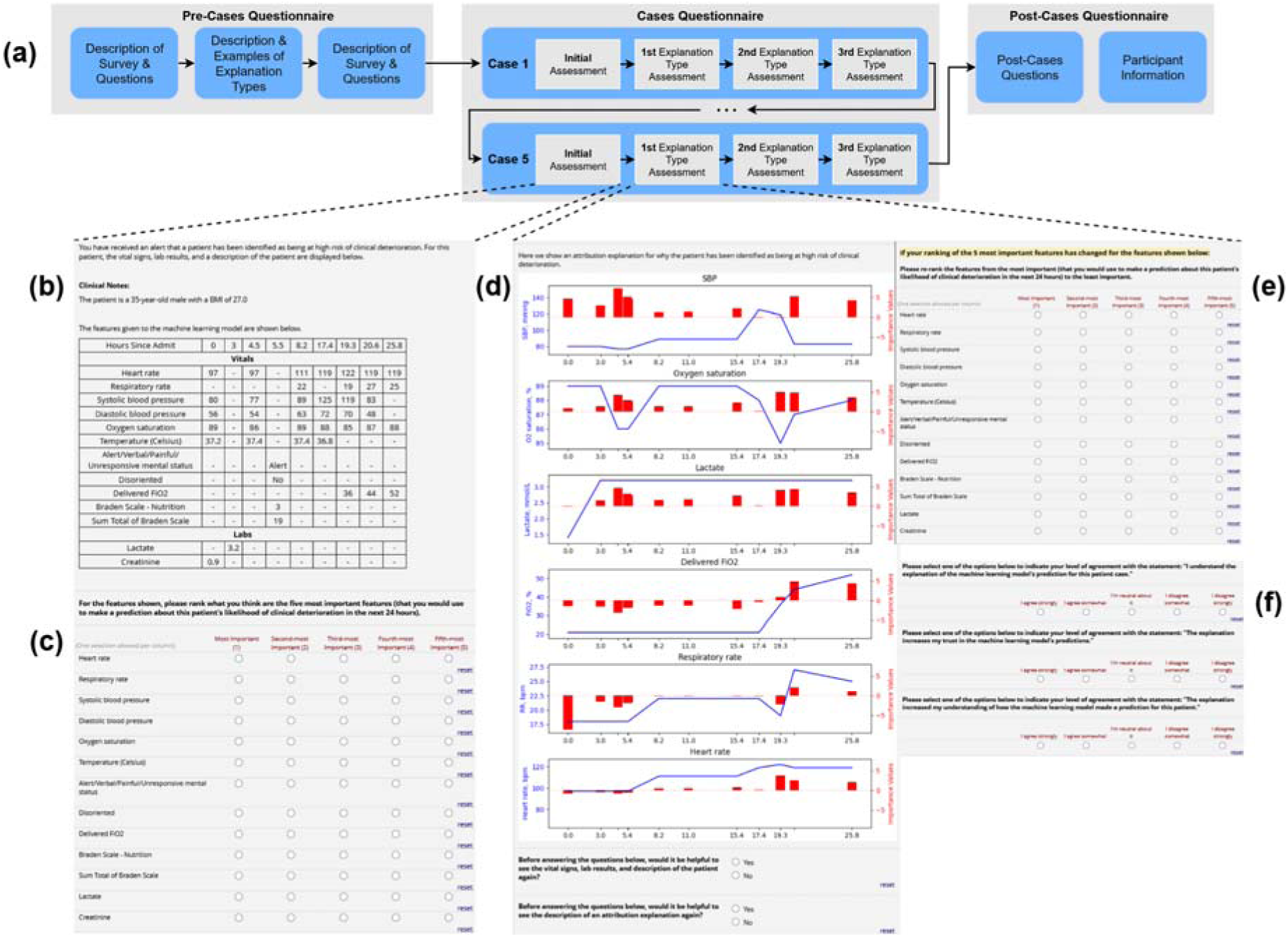
User study overview. (a) Participants are asked to review the user study task and explanation types and complete a Pre-Cases questionnaire, assess five distinct cases with explanations in the Cases questionnaire, and answer concluding questions about each explanation type in the Post-Cases questionnaire. In the Cases questionnaire, (b) participants see the patient’s vital, lab, and demographic features in the initial assessment, and (c) provide their initial ranking selection of what they think the most important features are. (d) For each explanation type assessment, participants see one of the three explanation types with the option to review the patient features and are prompted to (e) optionally revise their ranking and (f) answer explanation-specific questions.

### Pre-Cases Questionnaire

We provided participants with a description of the survey, details on the machine learning model’s prediction task, and an overview of the questions they would be asked. For each of the three explanation types, we described how the explanation could be interpreted and showed an example of the explanation. Then, we asked participants to rate how important they considered explanations and their trust in clinical ML models using a 5-point Likert scale [38]. We mapped Likert scale response options to numeric values. The response options and mappings were: *I agree strongly* (+2), *I agree somewhat* (+1), *I’m neutral about it* (0), *I disagree somewhat* (-1), *I disagree strongly* (-2).

### Cases Questionnaire

We showed each participant five patient cases in the Cases questionnaire. The process for a single case is as follows:

First, we informed participants that the ML model identified the patient as being at high risk of clinical deterioration. We asked participants to consider a table of patient feature values over time (Figure 3b) that displayed 13 out of the 55 features used by the model to make predictions. We included a feature in the table if it was shown as part of one of the explanations used in the user study. After showing the patient data table, we asked participants for their *initial ranking* of the five most important features that they think indicate that the patient is at high risk of clinical deterioration (Figure 3c).

Next, we showed participants explanations of each type (Figure 3d), one at a time, with the option to review the patient data table or the description of the explanation type. We showed explanations in random order for each case to prevent ordering effects from biasing results towards a specific explanation type. After showing each explanation, we asked participants to re-rank their selection of the top-5 features if the explanation affected the features they considered important (Figure 3e). Each re-ranking was pre-populated with the participant’s selections from the previous ranking for the same case. For example, the first explanation shown for case 2 was pre-populated with the participants’ selections in the initial ranking for case 2. We also asked participants to rate their agreement on a Likert scale with statements measuring increases in model trust and model understanding caused by the explanation, and if they understood the explanation (Figure 3f).

### Post-Cases Questionnaire

In the last section of the survey, we asked participants to rate their agreement on a Likert scale with statements measuring increases in model trust and understanding and changes to explanation understanding caused by each type of explanation in general. We also asked participants about their preferences for viewing multiple explanation types together and whether they favored shorter, more concise over longer, more detailed attribution and rule-based explanations. At the end of the questionnaire, we provided participants with a free-text response section to describe what they liked or disliked about each explanation type.

### Statistical Analysis

We used a one-sample Wilcoxon signed-rank test with a significance threshold of 95% to determine if each Likert scale question had a median response greater than zero.

We used Linear Mixed-effect Models (LMMs) to determine the extent to which variables of interest account for the survey responses. LMMs are an extension of simple linear models, incorporating both fixed effects and random effects [39]. We treated Participant ID as a random effect to account for variations in participants’ baseline values.

For each of the explanation importance, model trust, model understanding, explanation understanding, or influence dimensions, we fit two LMM models to participant responses. The first LMM was fit to participants’ responses from the Cases questionnaire with the explanation type, order in which the explanation was shown, case number, and participant’s role, recruitment site, gender, age, Pre-Cases questionnaire level of model trust, and Pre-Cases questionnaire level of explanation importance as covariates. The second LMM was fit to participants’ responses from the Post-Cases questionnaire with the explanation type and participant’s role, recruitment site, gender, age, Pre-Cases questionnaire level of model trust, and Pre-Cases questionnaire level of explanation importance as covariates.

For all LMMs, the categorical variables representing the explanation type, order in which the explanation was shown, case number, role, recruitment site, gender, and age were dummy encoded relative to a baseline category.

The coefficients assigned to each input variable by an LMM represent the effect size of the variable on the outcome. We used a Z-test and confidence threshold of 95% to determine if there was enough support to conclude that each coefficient (effect) was non-zero and that the corresponding variable had a statistically significant effect on the outcome.

We performed sentiment analysis on the free-text responses to evaluate whether participants considered explanations of each type to be *positive*, *negative*, or *both* positive and negative. Responses that did not contain positive or negative sentiments were considered *uncertain*.

## Results

We organize the results of our study by first considering explanation importance and then the four key dimensions of model trust, model understanding, explanation understanding, and influence. Lastly, we report survey responses pertaining to participants’ preferences for explanation representation.

### Explanation Importance

A clinician’s *explanation importance* refers to how important the clinician considers explanations of a clinical ML model’s predictions. Figure 4 displays these responses separated by participants’ roles, and Table 2 provides summary metrics for the responses separated by participants’ roles.

**Figure 4.**
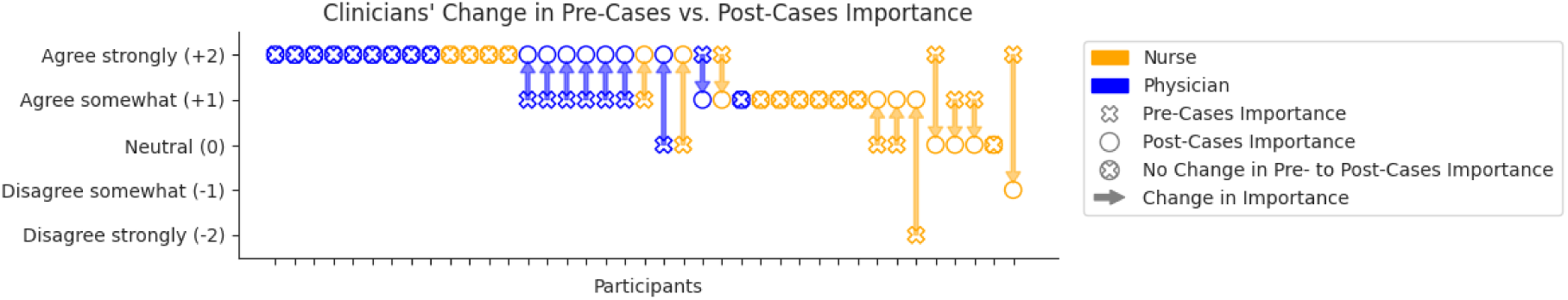
Participants’ assessment of the importance of having explanations. An -like symbol represents explanation importance responses from the Pre-Cases questionnaire, represents responses from the Post-Cases questionnaire, a -like symbol represents responses that were the same in the Pre- and the Post-Cases questionnaire, and arrows indicate the change from Pre-Cases to Post-Cases explanation importance.

**Table 2.** Explanation importance measured in the Pre-Cases and Post-Cases questionnaires.^a^.

| | Pre-Cases<br>Importance | Post-Cases<br>Importance | $p$-value |
| --- | --- | --- | --- |
| Nurses | $1.0 \pm 1.0$ | $1.0 \pm 0.84$ | .50 |
| Physicians | $1.50 \pm 0.62$ | $1.89 \pm 0.32$ | .017 |
| Combined | $1.23 \pm 0.87$ | $1.41 \pm 0.76$ | .127 |

<sup>a</sup>Numeric values are mapped from the Likert scale responses per the mapping shown in (Figure 4).

#### Explanations are important to participants

We found that average explanation importance was high in the Pre-Cases questionnaire and increased slightly in the Post-Cases questionnaire after participants saw all the explanations. However, we did not find a statistically significant difference between the Pre-Cases and Post-Cases responses for the combined results of nurses and physicians with a paired Wilcoxon signed-rank test (*P =*).

#### Seeing explanations affects participants’ explanation importance differently depending on role

When comparing Pre-Cases and Post-Cases measurements and separating results by role, we found that explanations affected nurses and physicians differently. A paired Wilcoxon signed-rank test revealed a significant increase in physicians’ explanation importance (*P =*) from the Pre-Cases to the Post-Cases questionnaire, while for nurses, explanation importance did not change (*P =*). These results indicate that physicians value explanations more after gaining experience with explanations, while nurses do not appear to change how they value explanations.

### Model Trust

A clinician’s *model trust* refers to the self-reported level of trust in predictions made by a model. In the Pre-Cases questionnaire, we measured baseline levels of trust in ML models (*initial trust*) by asking participants to rate their agreement that they trust the predictions of ML models in general. In the Cases questionnaire, participants provided ratings for each specific case and explanation pair, while in the Post-Cases questionnaire, participants provided ratings for each explanation type.

#### Explanations increase participants’ model trust

In the Pre-Cases questionnaire, participants reported average agreement on a 5-point Likert scale, with zero representing neutral, of 0.51 ± 0.82 (*P <* . 001), indicating that at baseline, participants were more trusting of clinical ML models than not. After being exposed to explanations, participants reported that explanations increased their trust in the predictions made by the ML model. Agreement for the increase in trust was 0.40 ± 0.92 (*P <* . 001) in the Cases and 0.54 ± 1.0 (*P <* . 001) in the Post-Cases questionnaires.

#### Attribution explanations increase model trust the most

Figure 5a shows each participant’s responses in the Cases questionnaire split by explanation type. Participants reported an average agreement of 0.54 ± 0.88 (*P <* . 001) for attribution, 0.37 ± 0.82 (*P <* . 001) for counterfactual, and 0.30 ± 1.03 (*P <* . 001) for rule-based explanations that the explanations increased their trust in the predictions made by the ML model. We analyzed participants’ Cases questionnaire responses with an LMM, adjusting for the order in which the explanations were shown, the case, and participants’ role, recruitment site, gender, age, initial trust, and initial importance to isolate the effect of counterfactual and rule-based explanations on increasing participants’ model trust relative to attribution explanations. We used Z-tests with a 95% confidence interval to determine if effect sizes were non-zero. We found that attribution explanations increased model trust by 0.17 ± 0.06 (*P =* . 007) relative to counterfactual and 0.21 ± 0.06 (*P =* . 001) relative to rule-based explanations. We did not find that any other covariates in the Cases LMM had statistically significant effects on model trust.

**Figure 5.**
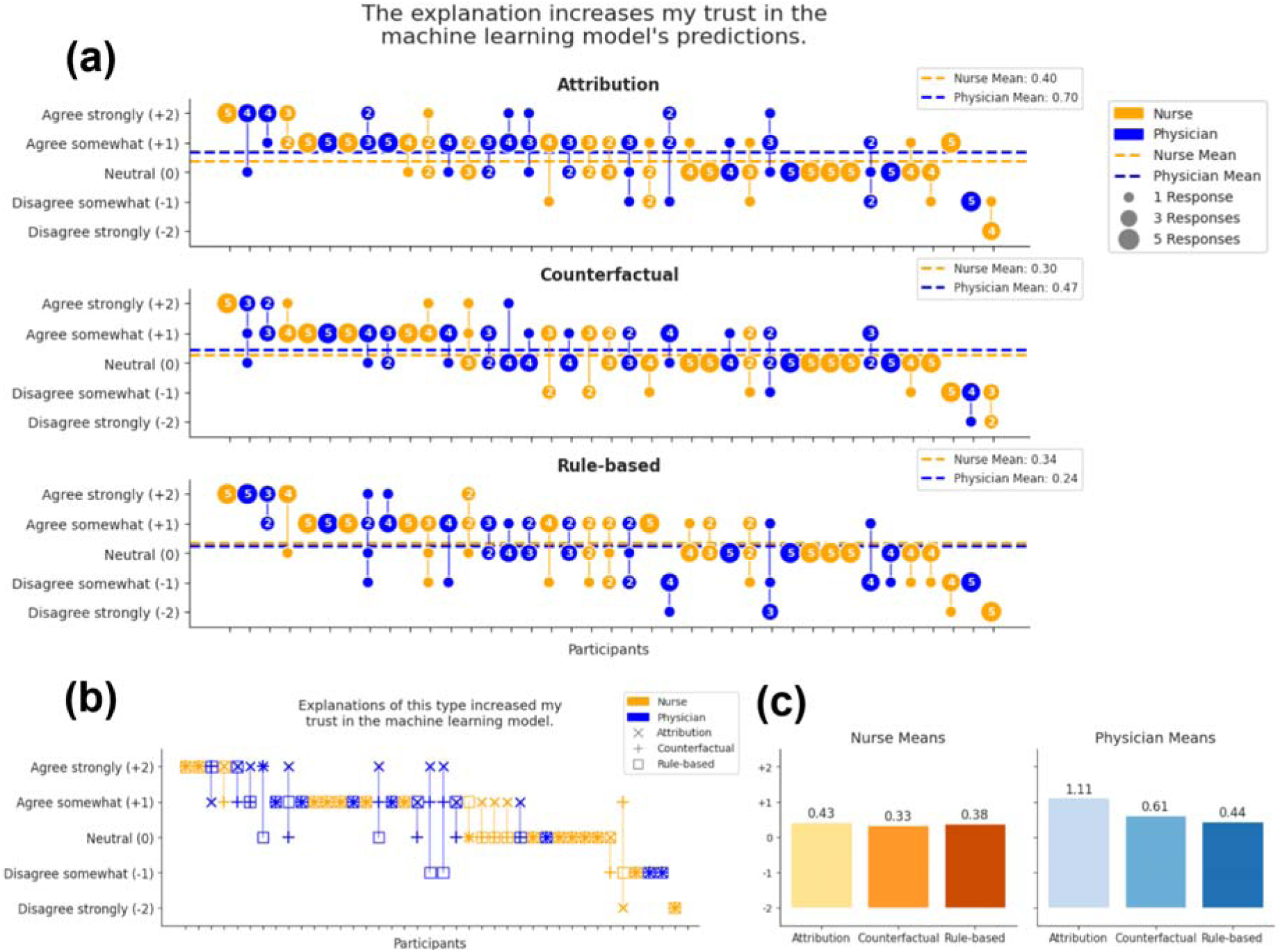
Participants’ Cases and Post-Cases responses by role and explanation type indicating agreement that the explanations increased the participants’ model trust. (a) Participants’ Cases responses. Each point represents a single response, with a vertical chain of points representing all of the responses for an individual participant. The size of and number inside each point represent the count of explanations for which the participant reported the same level of model trust. Horizontal lines represent the mean for each role. Participants are sorted from highest to lowest mean response value. (b) Post-Cases responses. For an individual participant, responses are oriented vertically and connected with a line. Each symbol indicates the response for a different explanation type. Participants are sorted from highest to lowest mean model trust across the three explanation types. (c) Bar plots of the mean Post-Cases model trust by role and explanation type. The number above each bar indicates the mean value.

Figure 5b shows each participant’s responses in the Post-Cases questionnaire for each explanation type. Participants reported an average agreement of 0.74 ± 1.09 (*P <* . 001) for attribution, 0.46 ± 0.91 (*P =* . 002) for counterfactual, and 0.41 ± 0.99 (*P =* . 008) for rule-based explanations that explanations of that type increased their model trust. Similar to the Cases response analysis, we used an LMM to analyze participants’ Post-Cases responses. We found that attribution explanations led to the greatest increase in participants’ model trust compared to counterfactual (0.24 ± 0.13, *P =* . 059) and rule-based (0.24 ± 0.13, *P =* . 059), although the difference between explanation types was not significant. We did not find that any other covariates in the Post-Cases LMM had statistically significant effects on model trust.

### Model Understanding

A clinician’s *model understanding* refers to the self-reported understanding of how the ML model makes predictions. In the Cases questionnaire, participants provided ratings for each specific case and explanation pair, while in the Post-Cases questionnaire, participants provided ratings for each explanation type.

#### Explanations increase participants’ model understanding

Participants reported an average agreement of 0.60 ± 0.96 (*P <* . 001) in the Cases questionnaire and 0.67 ± 1.08 (*P <* . 001) in the Post-Cases questionnaire, indicating that clinicians found that explanations increased their understanding of how the ML model made predictions in general.

#### Attribution explanations increase model understanding the most

Figure 6a shows each participant’s responses in the Cases questionnaire split by explanation type. Participants reported the highest mean agreement that the explanations increased model understanding for attribution explanations (0.74 ± 0.91), followed by counterfactuals (0.55 ± 0.91) and rule-based explanations (0.52 ± 1.05). We analyzed participants’ Cases questionnaire model understanding responses with an LMM and Z-tests and found that attribution explanations increased model understanding by 0.18 ± 0.07 (*P =* . 010) relative to counterfactual and 0.18 ± 0.07 (*P =* . 009) relative to rule-based explanations. We also found that the specific case had a significant effect on model understanding. Specifically, model understanding was lower by 0.20 ± 0.09 on average for Case 3 compared to Case 1.

**Figure 6.**
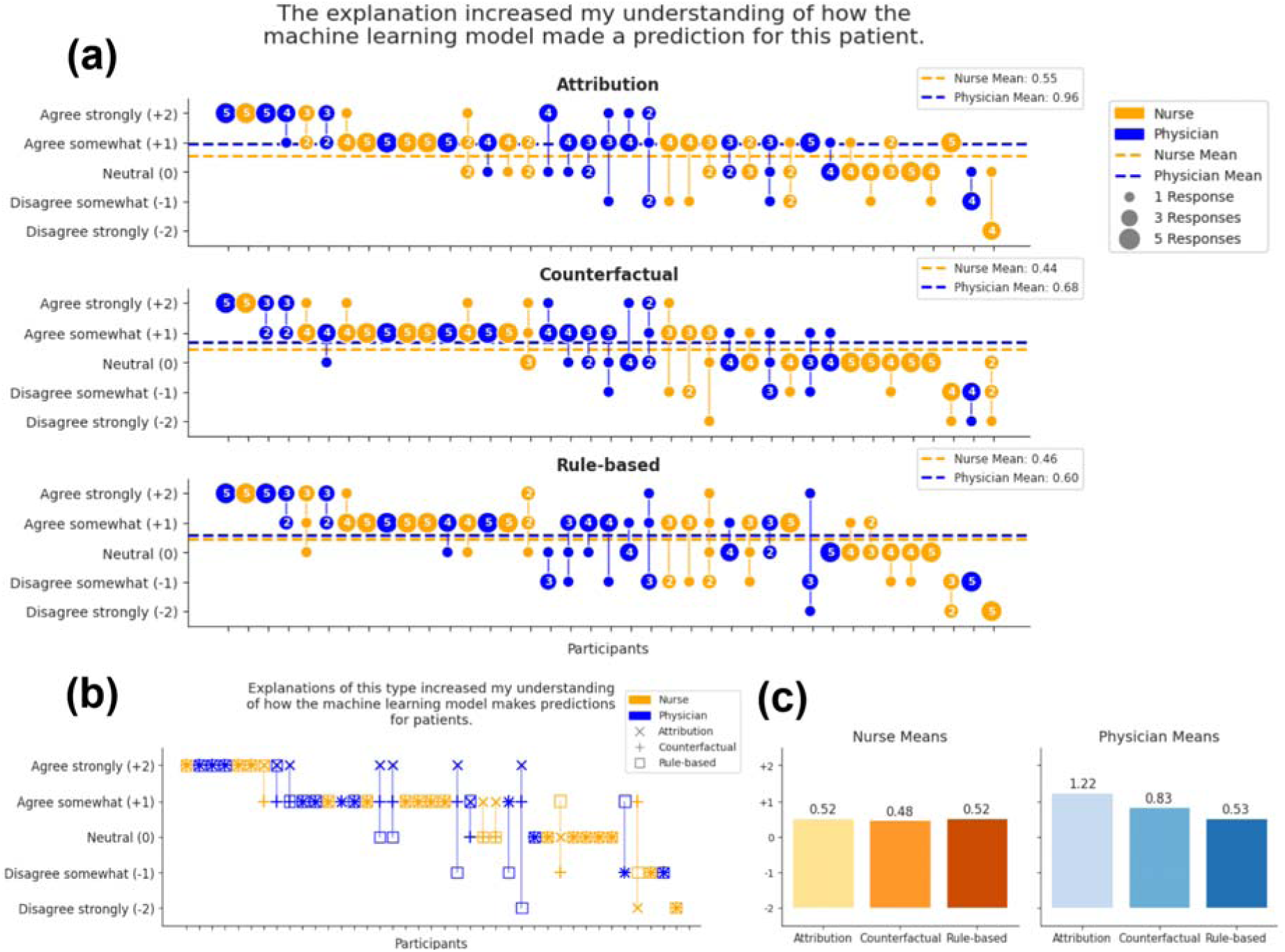
Participants’ Cases and Post-Cases responses by role and explanation type indicating agreement that the explanations increased the participants’ model understanding. (a) Participants’ Cases responses. Each point represents a single response, with a vertical chain of points representing all of the responses for an individual participant. The size of and number inside each point represent the count of explanations for which the participant reported the same level of model understanding. Horizontal lines represent the mean for each role. Participants are sorted from highest to lowest mean response value. (b) Post-Cases responses. Responses for an individual participant are oriented vertically and connected with a line. Each symbol indicates the response for a different explanation type. Participants are sorted from highest to lowest mean model understanding across the three explanation types. (c) Bar plots of the mean Post-Cases model understanding by role and explanation type. The number above each bar indicates the mean value.

Figure 6b shows each participant’s responses in the Post-Cases questionnaire for each explanation type. Participants reported an average agreement that the explanations increased their model understanding of 0.85 ± 1.14 (*P <* . 001) for attribution, 0.64 ± 0.96 (*P <* . 001) for counterfactual, and 0.53 ± 1.13 (*P =* . 005) for rule-based explanations. As with model trust, we analyzed participants’ Post-Cases questionnaire model understanding responses with an LMM and Z-tests and found that the difference between explanation types was not significant. We did not find that any other covariates in the Post-Cases LMM had statistically significant effects on model understanding.

### Explanation Understanding

In contrast to understanding how the model makes predictions (model understanding), *explanation understanding* refers to a participant’s self-reported understanding of a given explanation. In the Cases questionnaire, participants provided ratings for each specific case and explanation pair, while in the Post-Cases questionnaire, participants provided ratings for each explanation type.

#### Participants report understanding all explanation types

Participants reported an average agreement of 0.74 ± 0.97 (*P <* . 001) in the Cases questionnaire and 0.74 ± 1.09 (*P <* . 001) in the Post-Cases questionnaire, indicating that, in general, participants found explanations understandable.

#### Attribution explanations are the easiest type to understand

Figure 7a shows each participant’s responses in the Cases questionnaire split by explanation type. Participants reported that attribution explanations were the easiest to understand on average (0.88 ± 0.87), followed by counterfactuals (0.67 ± 0.93) and rule-based explanations (0.66 ± 1.09).

**Figure 7.**
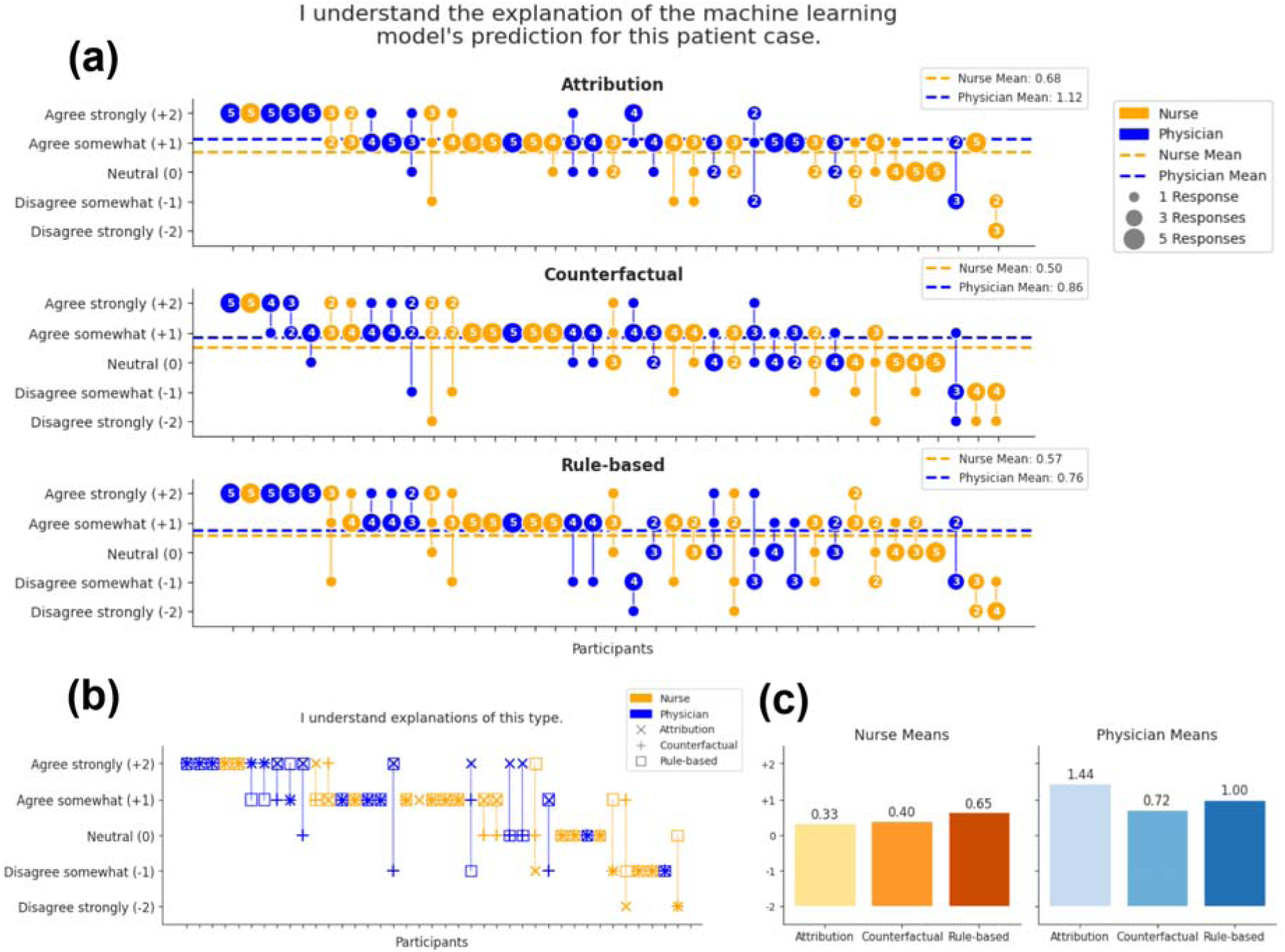
Participants’ Cases and Post-Cases explanation understanding by role and explanation type. (a) Participants’ Cases responses. Each point represents a single response, with a vertical chain of points representing all of the responses for an individual participant. The size of and number inside each point represent the count of explanations for which the participant reported the same level of explanation understanding. Horizontal lines represent the mean for each role. Participants are sorted from highest to lowest mean response value. (b) Post-Cases responses. Responses for an individual participant are oriented vertically and connected with a line. Each symbol indicates the response for a different explanation type. Participants are sorted from highest to lowest mean explanation understanding across the three explanation types. (c) Bar plots of the mean Post-Cases explanation understanding by role and explanation type. The number above each bar indicates the mean value.

We analyzed participants’ Cases questionnaire explanation understanding responses with an LMM and Z-tests. We found that participants reported attribution explanations were easier to understand by 0.23 ± 0.07 (*P =* . 001) relative to counterfactual and 0.22 ± 0.07 (*P =* . 003) relative to rule-based explanations. The case and participants’ initial trust in ML models also contributed to the explanation understanding. Explanations were more difficult to understand for Case 3 compared to Case 1, averaging 0.24 ± 0.09 (*P =* . 013) lower response values. Participants with higher initial trust found explanations easier to understand; every additional point of initial trust increased explanation understanding by 0.34 ± 0.17 (*P =* . 047).

Figure 7b shows each participant’s responses in the Post-Cases questionnaire for each explanation type. Participants reported an average explanation understanding of 0.85 ± 1.20 (*P <* . 001) for attribution, 0.55 ± 1.08 (*P =* . 002) for counterfactual, and 0.82 ± 0.98 (*P <* . 001) for rule-based explanations. An LMM analysis of these results indicated that the difference between explanation types was not significant. We did not find that any other covariates in the Post-Cases LMM had statistically significant effects on model understanding.

### Influence

We define the *influence* of an explanation as the effect of the explanation on a participant’s assessment of what features are most important for determining a patient’s risk of clinical deterioration. We measured influence in two ways. First, through the *Count of Rank Changes* (CRC) metric calculated over participants’ ranking selections in the Cases questionnaire. CRC quantifies the number of features whose rankings participants change after seeing an explanation. Second, through Likert scale responses to one question in the Post-Cases questionnaire, in which participants reported how much they felt explanations of each type influenced the features that they thought were most important for determining a patient’s risk of clinical deterioration (*self-reported influence*).

#### Explanations influence which factors participants consider important for determining a patient’s status

For CRC in the Cases questionnaire, the average was 0.68 ± 1.30 (*P <* . 001) ranking changes after seeing each explanation. For the Likert-scale responses in the Post-Cases questionnaire, the average was 0.34 ± 1.24 (*P =* . 003) over all explanation types. These results indicate that explanations both led to tangible changes in the features that participants considered important for determining that patients were at high risk of deterioration and were perceived as influential by participants.

#### Attribution explanations are the most influential

Figure 8a shows each participant’s CRC for the Cases questionnaire split by explanation type. In the Cases questionnaire, attribution explanations led to the highest CRC (0.81 ± 1.44), followed by rule-based (0.68 ± 1.28) and counterfactual explanations (0.54 ± 1.17). We analyzed participants’ CRC in the Cases questionnaire with an LMM and Z-tests and found that CRC for attribution explanations was higher by 0.31 ± 0.11 (*P =* . 005) relative to counterfactual and 0.24 ± 0.11 (*P =* . 030) relative to rule-based explanations.

**Figure 8.**
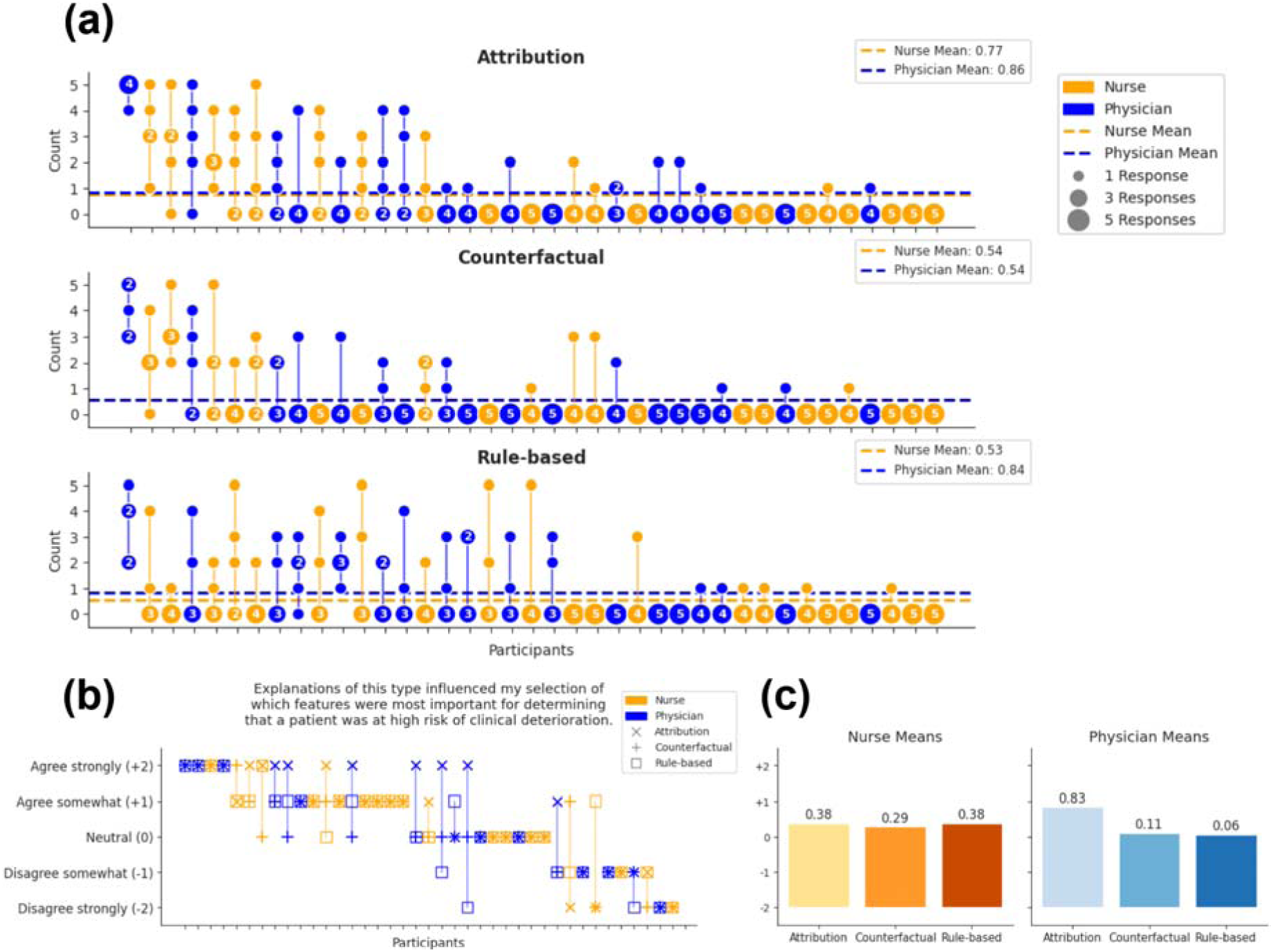
Participants’ Cases and Post-Cases counts of ranking changes by role and explanation type. (a) Participants’ Cases responses. Points represent ranking responses, with the corresponding y-axis value representing the number of features ranked differently compared to the previous ranking. The size of and number inside each point represent the number of explanations for which the participant had the same count of ranking changes value. Horizontal lines represent the mean for each role. Participants are sorted from highest to lowest mean count. (b) Post-Cases responses. Responses for an individual participant are oriented vertically and connected with a line. Each symbol indicates the response for a different explanation type. Participants are sorted from highest to lowest mean influence across the three explanation types. (c) Bar plots of the mean Post-Cases influence by role and explanation type. The number above each bar indicates the mean value.

We also found that the order in which explanations were shown and the case significantly affected the CRC in the Cases questionnaire. CRC was lower for explanations shown later in the ordering. Cases 2 through 5 had lower CRC than Case 1, although the difference varied by case. Tables showing the effects of the order in which the explanation was shown and the case on the CRC can be found in the supplementary materials. We did not find that any other covariates in the Cases LMM had statistically significant effects on CRC.

Figure 8b shows each participant’s self-reported influence in the Post-Cases questionnaire for each explanation type. Participants reported an average influence of 0.59 ± 1.35 (*P =* . 007) for attribution, 0.21 ± 1.15 (*P =* . 16) for counterfactual, and 0.23 ± 1.20 (*P =* . 13) for rule-based explanations. Analysis of participants’ Post-Cases influence with an LMM and Z-tests showed that attribution explanations were more influential than counterfactuals by 0.32 ± 0.15 (*P =* . 034) and rule-based by 0.35 ± 0.15 (*P =* . 021) explanations. We did not find that any other covariates in the Post-Cases LMM had statistically significant effects on self-reported influence.

### Preferences for Explanation Representation

In the Post-Cases questionnaire, we asked participants about their preferences for seeing multiple explanation types together and if they would prefer to see more concise or more detailed attribution and rule-based explanations. We also gave participants the option to specify what they liked or disliked about each type of explanation with short free-text responses.

Figure 9 shows participants’ preferences for seeing multiple types of explanations together (a), and for those who said they wanted to see multiple types together, shows the combinations of types they selected (b). Figure 9 also shows participants’ preferences for seeing shorter or longer attribution (c) and rule-based (d) explanations. Table 3 shows participants’ free-text responses, coded by sentiment towards the explanation type.

**Figure 9.**
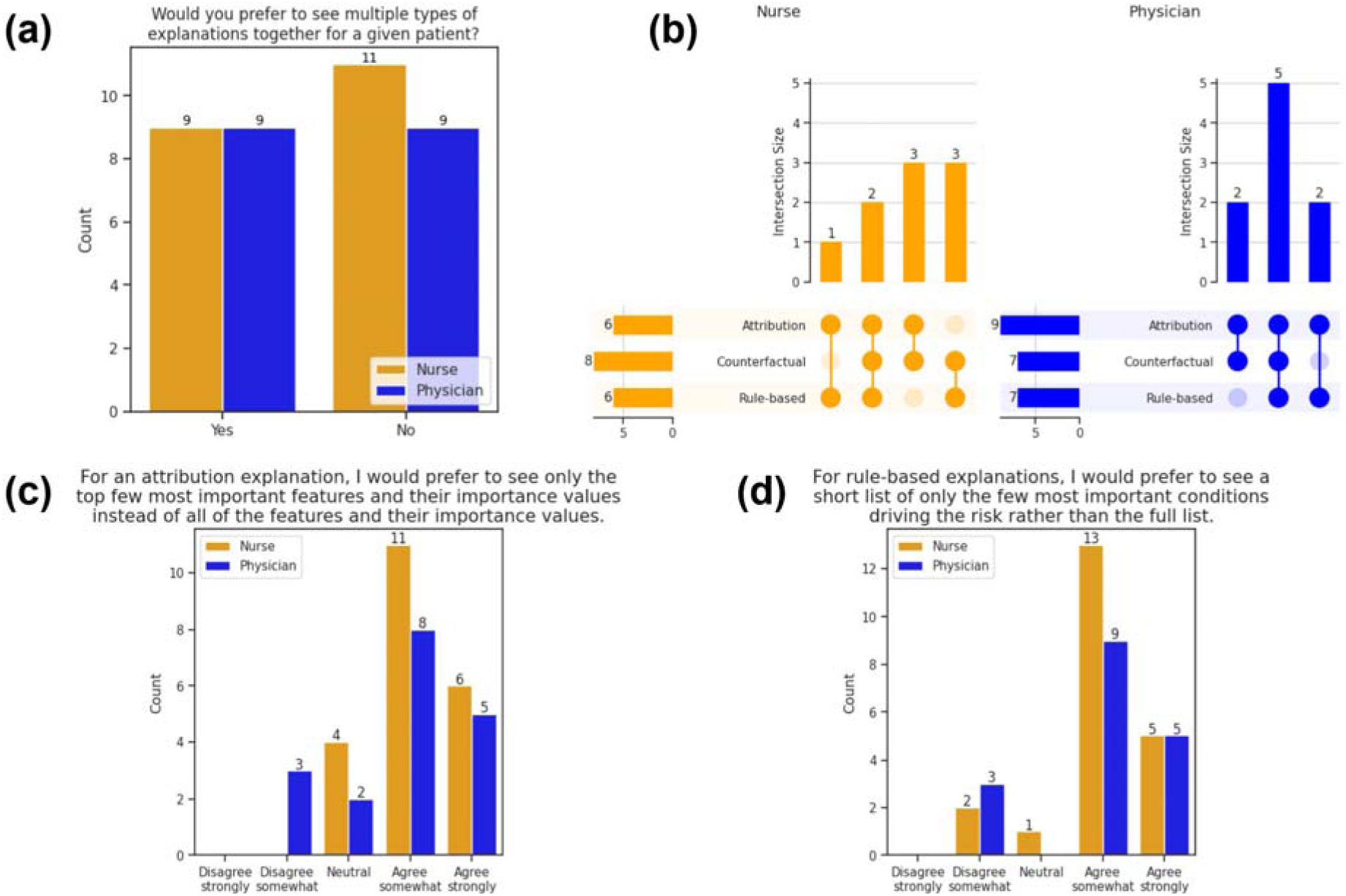
Participants’ preferences for how explanations should be represented. (a) Counts of participants who indicated that they do or do not want to see more than one type of explanation together. For the 18 total participants who reported that they wanted to see multiple explanation types together, we show (b) Upset plots indicating the combinations of explanation types that nurses and physicians prefer to see together. (c) Participants’ preferences for more concise or more detailed attribution explanations. (d) Participants’ preferences for more concise or more detailed rule-based explanations.

**Table 3.** Participants’ free-text response sentiments by explanation type.^a^.

|  | Positive | Negative | Both | Uncertain |
| --- | --- | --- | --- | --- |
| Attribution | 15 (62.5 %) | 5 (20.8 %) | 2 (8.3 %) | 2 (8.3 %) |
| Counterfactual | 4 (17.4 %) | 15 (65.2 %) | 4 (17.4 %) | 0 (0.0 %) |
| Rule-based | 5 (20.0 %) | 12 (48.0 %) | 6 (24.0 %) | 2 (8.0 %) |

<sup>a</sup>We labeled responses as positive or negative depending on what the response said about the explanation type. *Both* responses included both positive and negative sentiments. *Uncertain* comments did not contain positive or negative sentiments about the explanation type.

#### Participants want to see multiple types of explanations together

46.2% (18/39) of participants report wanting to see multiple explanation types together, and of those 18 responses, 38.8% (7/18) wanted to see all three types of explanations together. Attribution and counterfactual explanations appeared equally as often (15/18, 83.3%) in the combinations of multiple types that participants indicated they wanted to see. Rule-based explanations were selected the least often (13/18, 72.2%).

#### Participants prefer more concise attribution and rule-based explanations over more detailed explanations

For attribution explanations, 76.9% (30/39) of participants reported wanting to see the top few most important features and their importance values rather than seeing all of the features and their importance values. For rule-based explanations, 84.2% (32/38) of participants reported wanting to see a short list of only the few most important conditions driving the risk rather than seeing the full list of conditions. One participant did not provide a response.

#### Participants like attribution explanations the most

Overall, participants responded with substantially more positive comments for attribution compared to counterfactual or rule-based explanations in the free-text responses, indicating that participants felt most positively towards attribution explanations. Although rule-based explanations had more positive comments compared to counterfactuals, both rule-based and counterfactual explanations had similar distributions of responses.

## Discussion

Our work aimed to compare the most prominent local post-hoc model-agnostic explanation types. We focused on a realistic clinical setting, using a risk prediction model trained on longitudinal patient data and real attribution, counterfactual, and rule-based methods to generate explanations. We compared the effects of different explanation types on how important clinicians consider explanations and the key dimensions of model trust, model understanding, explanation understanding, and influence.

### Explanations of all types are valued by clinicians

All three types of explanations show effects across the four key dimensions of model trust, model understanding, explanation understanding, and influence. Our findings that clinicians understand all three types of explanations and that all types increase model trust and model understanding suggest explanations can improve clinicians’ interaction with ML-based clinical decision support systems. Our results extend previous work showing that attribution and counterfactual explanations can increase users’ self-reported trust in and understanding of ML models and affect users’ decision-making, and that attribution explanations have greater impacts than counterfactual explanations [22]. Our findings that all three types of explanations influence clinicians’ understanding of the features important for determining a patient’s risk of clinical deterioration demonstrate that explanations can improve clinicians’ decision-making by highlighting important features and allowing clinicians to review a ‘second opinion’ through the explanation. However, explanations must be used cautiously, as previous work has shown that explanations can lead to poorer decisions when paired with incorrect model predictions [7,29].

### Attribution explanations have the most value to clinicians across all of the key dimensions measured

Based on our empirical findings, we recommend that clinical ML systems prioritize attribution-based explanations as the primary explanation modality, supplemented by counterfactual or rule-based explanations. Attribution explanations outperform counterfactual and rule-based explanations across the model trust, model understanding, explanation understanding, and influence dimensions, and were associated with more positive comments in the free-text responses than counterfactual or rule-based explanations. Our results showing that attribution explanations are valued more than other explanation types extend previous work that shows attribution explanations are more influential and affect trust and understanding more than counterfactual explanations [22,40].

### Nurses and physicians differ in how important they consider explanations, and this difference increases after seeing explanations

We hypothesize that the difference between nurses and physicians in initial explanation importance and the change in explanation importance induced by explanations is because explanations affect the workflows of nurses and physicians differently. Physicians’ workflows include more clinical decision-making, such as diagnosing patients and developing treatment plans. For these tasks, explanations may be beneficial by providing support for or helping clinicians reason through a decision. In contrast, nurses have a more protocolized workflow where explanations may not be as useful, as explanations are less likely to affect standardized protocols. This aligns with previous work by Barda et al. [41] that shows that physicians seek explanations that provide insights into patients’ health statuses, while nurses seek explanations that provide directly actionable information.

### Effective clinical ML model explanation systems should implement several different explanation types

Clinicians’ preferences for seeing multiple explanation types simultaneously suggest that different explanation types may serve complementary functions in clinical reasoning.

Our findings may be especially consequential for the specific predictive task that our study is grounded in. When clinicians are alerted that a particular patient is at risk of clinical deterioration, knowing *why* the model is identifying a patient as being at risk of clinical deterioration allows the clinicians to make more informed decisions about what to do next. Understanding how explanation types differ in value in this context can lead to changes in clinical decision-making that substantially impact patient outcomes.

There are several limitations to our study that should be kept in mind when interpreting the results. We used one representative method with default hyperparameter settings to generate explanations of each type. Additionally, we generated explanations for only one deep learning model and time series task. As such, our results may to some extent reflect limitations in the explanation methods used to generate explanations from time series data, rather than fundamental issues with the explanation types themselves. Explanations can vary substantially depending on the model and task [42,43] and can be sensitive to the hyperparameters used in the explanation-generating method [44,45]. However, to mitigate this limitation, we examined the explanations used to assess their face validity.

Clinicians’ responses to the different explanation types may depend on the visual format in which each explanation type is presented. To address this limitation, we used standard visual conventions for each type of explanation.

Additionally, our results for model trust, model understanding, explanation understanding, and Post-Cases influence are based on clinicians’ self-reported measures. Previous work has found that self-reported measures may not always align with objective measures [46,47].

Finally, our user study cohort of 39 clinicians represents a limited sample from two academic medical centers. As such, our results may not fully generalize to community hospitals or resource-constrained environments where clinicians have different baseline attitudes towards or familiarity with ML models. However, we note that our cohort represents one of the largest user studies of explanations involving clinicians.

In conclusion, our user study helps identify how explanation types differ in their value to clinicians. We find that attribution, counterfactual, and rule-based explanations affect model trust, model understanding, explanation understanding, and influence, with attribution explanations having the greatest effects across all measured dimensions. Our findings have important implications for the development and deployment of XAI methods for explaining machine learning models in clinical settings.

## Supporting information

Supplemental Survey Questions

## Data Availability

The data analyzed during the current study cannot be shared publicly due to restrictions in the data-use agreements with the two health systems from which the data was collected. The synthetic data cases and explanations generated are available in the GitHub repository [48].

## Authors’ Contributions

B.B. contributed to the conceptualization, data curation, formal analysis, methodology, visualization, software, and writing (original draft).

M.O. contributed to the investigation, project administration, and writing (review & editing).

O.T.N. and D.A.W. contributed to the conceptualization and writing (review & editing).

K.A.C. contributed to the data curation and writing (review & editing).

J.M. and C.A.K. contributed to the software and writing (review & editing).

M.A. and D.P.E. contributed to the investigation, project administration, and writing (review & editing).

A.M. contributed to the conceptualization, methodology, validation, funding acquisition, and writing (review & editing).

M.M.C. contributed to the conceptualization, methodology, validation, funding acquisition, and writing (review & editing).

M.C. contributed to the conceptualization, methodology, supervision, funding acquisition, and writing (review & editing).

