## Supplemental Tables 1 and 2 for "How Best to Explain Machine Learning Models to Clinicians: A User Study of Explanation Types"

### Supplementary Section 1 Additional Findings

To evaluate the effect of the case number, we used the LMM analysis to identify the effect sizes of each Case 2-5 relative to Case 1 on CRC. Table 1 shows the effect sizes of the case on CRC. The results indicate that for later questions in the survey, clinicians are less likely to choose different re-rankings of the important features.

Supplementary Table 1: Effect of the Case on the Count of Ranking Changes (CRC) value relative to the first case.

| Case Number | Relative CRC | $p$-value |
|---|---|---|
| Case 1 | - | - |
| Case 2 | $-0.63 \pm 0.14$ | $< 0.001$ |
| Case 3 | $-0.44 \pm 0.14$ | 0.002 |
| Case 4 | $-0.86 \pm 0.14$ | $< 0.001$ |
| Case 5 | $-0.64 \pm 0.14$ | $< 0.001$ |

As with the case number, we used an LMM to analyze the order in which the explanations were shown. Table 2 shows the effects of the order in which explanations were shown on the CRC. The results indicate that explanations shown later led to fewer ranking changes than explanations that were shown first.

Supplementary Table 2: Effect of the order in which the explanation was shown on the Count of Ranking Changes (CRC) value relative to the first case.

| Explanation Order | Relative CRC | $p$-value |
|---|---|---|
| First | - | - |
| Second | $-0.47 \pm 0.11$ | $< 0.001$ |
| Third | $-0.52 \pm 0.11$ | $< 0.001$ |
