## Supplemental Survey Questions for "How Best to Explain Machine Learning Models to Clinicians: A User Study of Explanation Types"

### User Preferences for Machine Learning Model Explanations

Returning?

A A A

Page 1 of 26

**Researchers at UW-Madison and the University of Chicago have developed a set of algorithms that feed a new clinical decision support tool that automatically assesses hospitalized adult patient's risk for Clinical Deterioration and further recommends diagnoses and treatments for the clinical team to consider.**

**To gather feedback on how to best display and explain the algorithms to maximize your understanding, trust, and satisfaction with the predictions our models make (including what is driving the prediction), we are asking you to complete this short online research survey.**

**Please be sure to complete this survey on a computer**, as the questions and images contained in the survey have only been formatted and tested for use with a computer, and are not formatted for phones or tablets.

**Informed consent information:** The survey is **voluntary** and should take **less than 30 minutes** to complete. You will be compensated $50 for your time. You can skip any survey questions that you do not want to answer. Even if you start the survey, you are not required to complete it. You can stop at any time. We will ask you for some additional information about yourself at the end of the survey in order to pay you for your time and fulfill reporting requirements. All of your answers will be confidential. We will keep all respondent-level survey data here at UW Madison, but may share summary data, selected free-text responses, and findings (all de-identified) with our collaborating teams at the University of Chicago, and partnering company AgileMD, Inc. (the group helping us design the clinical decision support tool interface). This shared data and the results of this research study may be published in a journal for the purpose of advancing medical knowledge. You will not be identified by name or by any other identifying information in any publication or report about this research.

If you have any questions about the survey, clinical decision support tool, or the study overall, please contact Matthew Churpek and program manager Madeline Oguss at.

This research was funded by The National Heart, Lung, and Blood Institute (NHLBI) grant number R01-HL157262.

Thank you in advance for your feedback!

Next Page >>

Save & Return Later

Powered by REDCap

### User Preferences for Machine Learning Model Explanations

A A A

Page 2 of 26

### Overview of the Survey

**The task:**

Our main set of questions will ask you to evaluate the importance of different *features* used to determine that a given patient in the medical-surgical wards is at high risk of clinical deterioration. A feature is an individual patient characteristic, lab, or vital sign value, such as "Heart rate" or "Respiratory rate" that might influence the prediction that a patient is at high risk of clinical deterioration.

For each patient predicted to be high-risk, you will be presented with the patient's features. First, you will be asked to select and rank the five most important features for determining that the patient is at high risk of clinical deterioration.

Next, you will be shown one of three explanations for why the machine learning model predicted that the patient was at high risk. After you evaluate the explanation, you will be asked if the explanation has affected the features you consider to be important for determining that the patient is at high risk of clinical deterioration and you will be allowed to re-rank the features.

Two more explanations of different types will be shown with the same format as above. If your ranking changes after seeing an explanation, you will have the option to re-rank the feature values.

We will also ask you a few introductory questions before and a few concluding questions after you complete the main task.

**What does the machine learning model do (what type of prediction/output is it making)?**

The machine learning model in this survey uses patient data to predict whether a given patient in the medical-surgical wards is at high risk of clinical deterioration in the next 24 hours.

**What is an explanation of the machine learning model's prediction?**

An explanation characterizes the features that were most important for the prediction made by a machine learning model. We will show you three different types of explanations.

**You will see three different types of explanations: attribution explanations, counterfactual explanations, and rule-based explanations. Each method uses a different representation to indicate how the model is using a patient's features to predict the patient's risk of clinical deterioration.**

**What is an attribution explanation?**

An attribution explanation provides an 'importance' value for each feature that represents the extent to which the feature contributed to the model's prediction. A positive importance value indicates that the feature at a given time point influences the model to predict that the patient is at high risk of clinical deterioration. In contrast, a negative importance value indicates that the feature at a given time point influences the model to predict that the patient is at low risk of clinical deterioration.

For example, in the attribution explanation below, the blue line denotes the heart rate measurements through time and the red bars denote the importance values for the heart rate feature for times between 8.4 and 38.9 hours after admission. According to the model, the heart rate values between hours 8.4 - 36.2 slightly decrease the patient's risk, and the values at hours 36.2 - 38.9 significantly increase the patient's risk.

**Example explanation:**

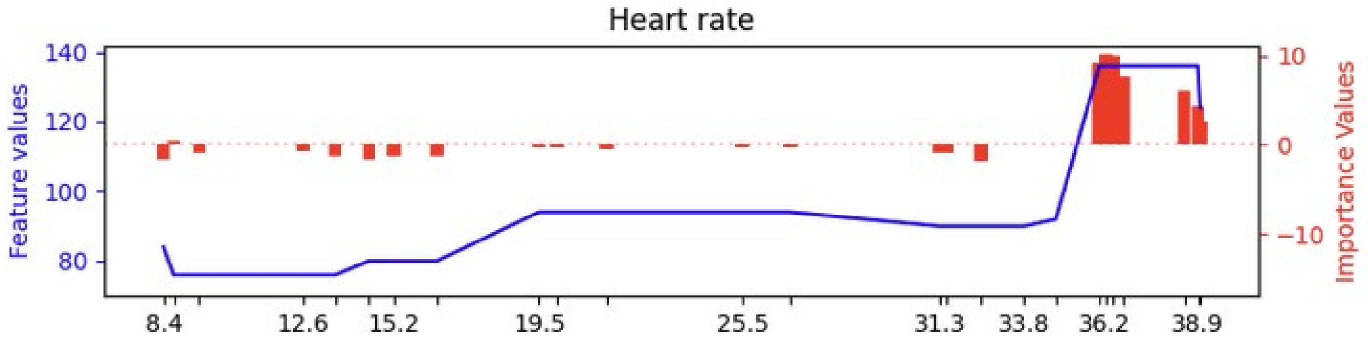


**What is a counterfactual explanation?**

A counterfactual explanation asks "What would need to change about the patient's feature values to change the prediction from high risk to no longer high risk?"

For example, for a patient who is predicted to be at high risk of clinical deterioration, the counterfactual explanation may tell us that if we replace the heart rate values for the patient (where the blue line denotes the heart rate measurements through time) with a sequence of measurements that is lower (the red dotted line), it will change the model's prediction to say the patient is instead at low risk of clinical deterioration. Effectively, the counterfactual explanation shows us a minimum-size subset of features that can be replaced to change the prediction from high risk of clinical deterioration to low risk of clinical deterioration.

**Example explanation:**

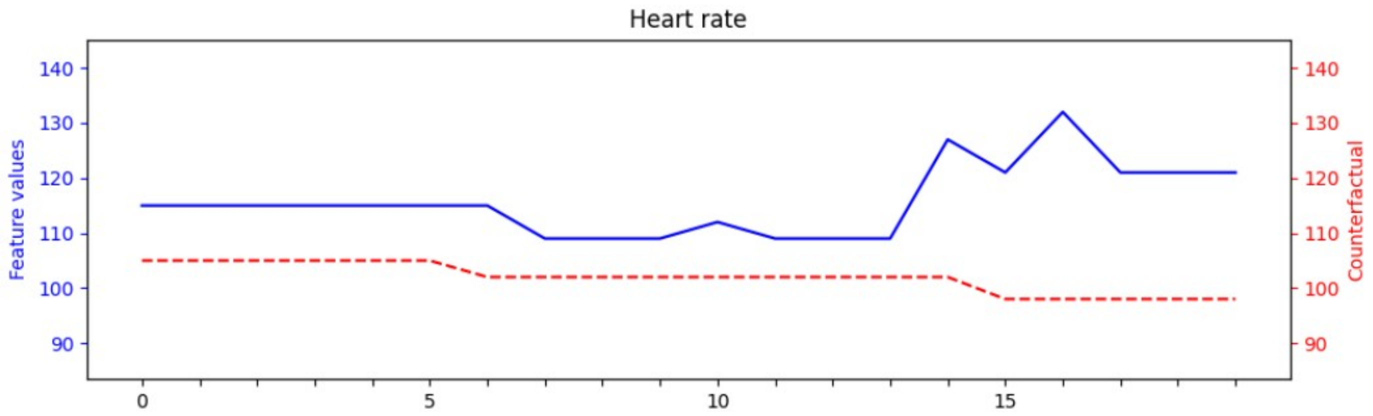


**What is a rule-based explanation?**

A rule-based explanation tells us combinations of feature values that lead the model to predict the patient's risk of clinical deterioration. The explanation is formulated as a rule in which each part of the rule compares a feature to a specific value or threshold.

For example, given a patient is predicted to be at high risk of clinical deterioration, the rule-based explanation might posit that the risk is due to both the Heart rate at a specific time being over a threshold and the Respiratory rate at a specific time being over a threshold.

**Example Explanation:**

**BECAUSE:**

Heart rate at time 36.2 > 110

Respiratory rate at time 39.8 > 25

**THEN:**

This patient is expected to be at high risk of clinical deterioration.

<< Previous Page

Next Page >>

Save & Return Later

Powered by REDCap

### User Preferences for Machine Learning Model Explanations

A A A

Page 3 of 26

### Introductory Questions

**Please select one of the options below to indicate your level of agreement with the statement: "It is important to me to have an explanation of a machine learning model's predictions."**

| I agree strongly | I agree somewhat | I'm neutral about it | I disagree somewhat | I disagree strongly |
|---|---|---|---|---|
| ○ | ○ | ○ | ○ | ○ |

**Please select one of the options below to indicate your level of agreement with the statement: "In general, I trust the predictions of machine learning models in clinical settings."**

| I agree strongly | I agree somewhat | I'm neutral about it | I disagree somewhat | I disagree strongly |
|---|---|---|---|---|
| ○ | ○ | ○ | ○ | ○ |

**What information do you think the explanation for a machine learning model's prediction should convey?**

<< Previous Page

Next Page >>

Save & Return Later

Powered by REDCap

### User Preferences for Machine Learning Model Explanations

AAA

Page 4 of 26

### Survey Question 1

**You have received an alert that a patient has been identified as being at high risk of clinical deterioration. For this patient, the vital signs, lab results, and a description of the patient are displayed below.**

**Clinical Notes:**

The patient is a 35-year-old male with a BMI of 27.0

**The features given to the machine learning model are shown below.**

| Hours Since Admit | 0 | 3 | 4.5 | 5.5 | 8.2 | 17.4 | 19.3 | 20.6 | 25.8 |
|---|---|---|---|---|---|---|---|---|---|
| **Vitals** | | | | | | | | | |
| Heart rate | 97 | - | 97 | - | 111 | 119 | 122 | 119 | 119 |
| Respiratory rate | - | - | - | - | 22 | - | 19 | 27 | 25 |
| SBP | 80 | - | 77 | - | 89 | 125 | 119 | 83 | - |
| DBP | 56 | - | 54 | - | 63 | 72 | 70 | 48 | - |
| Oxygen saturation | 89 | - | 86 | - | 89 | 88 | 85 | 87 | 88 |
| Temperature (Celsius) | 37.2 | - | 37.4 | - | 37.4 | 36.8 | - | - | - |
| AVPU mental status | - | - | - | Alert | - | - | - | - | - |
| Disoriented | - | - | - | No | - | - | - | - | - |
| Delivered FiO2 | - | - | - | - | - | - | 36 | 44 | 52 |
| Braden Scale - Nutrition | - | - | - | 3 | - | - | - | - | - |
| Sum Total of Braden Scale | - | - | - | 19 | - | - | - | - | - |
| **Labs** | | | | | | | | | |
| Lactate | - | 3.2 | - | - | - | - | - | - | - |
| Creatinine | - | - | - | - | - | - | - | - | - |

**For the features shown, please rank what you think are the five most important features (that you would use to make a prediction about this patient's likelihood of clinical deterioration in the next 24 hours).**

| (One selection allowed per column) | Most Important (1) | Second-most Important (2) | Third-most Important (3) | Fourth-most Important (4) | Fifth-most Important (5) |
|---|---|---|---|---|---|
| **Heart rate** | ○ | ○ | ○ | ○ | ○ |
| **Respiratory rate** | ○ | ○ | ○ | ○ | ○ |
| **SBP** | ○ | ○ | ○ | ○ | ○ |
| **DBP** | ○ | ○ | ○ | ○ | ○ |
| **Oxygen saturation** | ○ | ○ | ○ | ○ | ○ |
| **Temperature (Celsius)** | ○ | ○ | ○ | ○ | ○ |
| **AVPU mental status** | ○ | ○ | ○ | ○ | ○ |
| **Disoriented** | ○ | ○ | ○ | ○ | ○ |
| **Delivered FiO2** | ○ | ○ | ○ | ○ | ○ |
| **Braden Scale - Nutrition** | ○ | ○ | ○ | ○ | ○ |
| **Sum Total of Braden Scale** | ○ | ○ | ○ | ○ | ○ |
| **Lactate** | ○ | ○ | ○ | ○ | ○ |
| **Creatinine** | ○ | ○ | ○ | ○ | ○ |

<< Previous Page

Next Page >>

Save & Return Later

Powered by REDCap

**User Preferences for Machine Learning Model Explanations**

AAA



### Survey Question 1

**Here we show an attribution explanation for why the patient has been identified as being at high risk of clinical deterioration.**

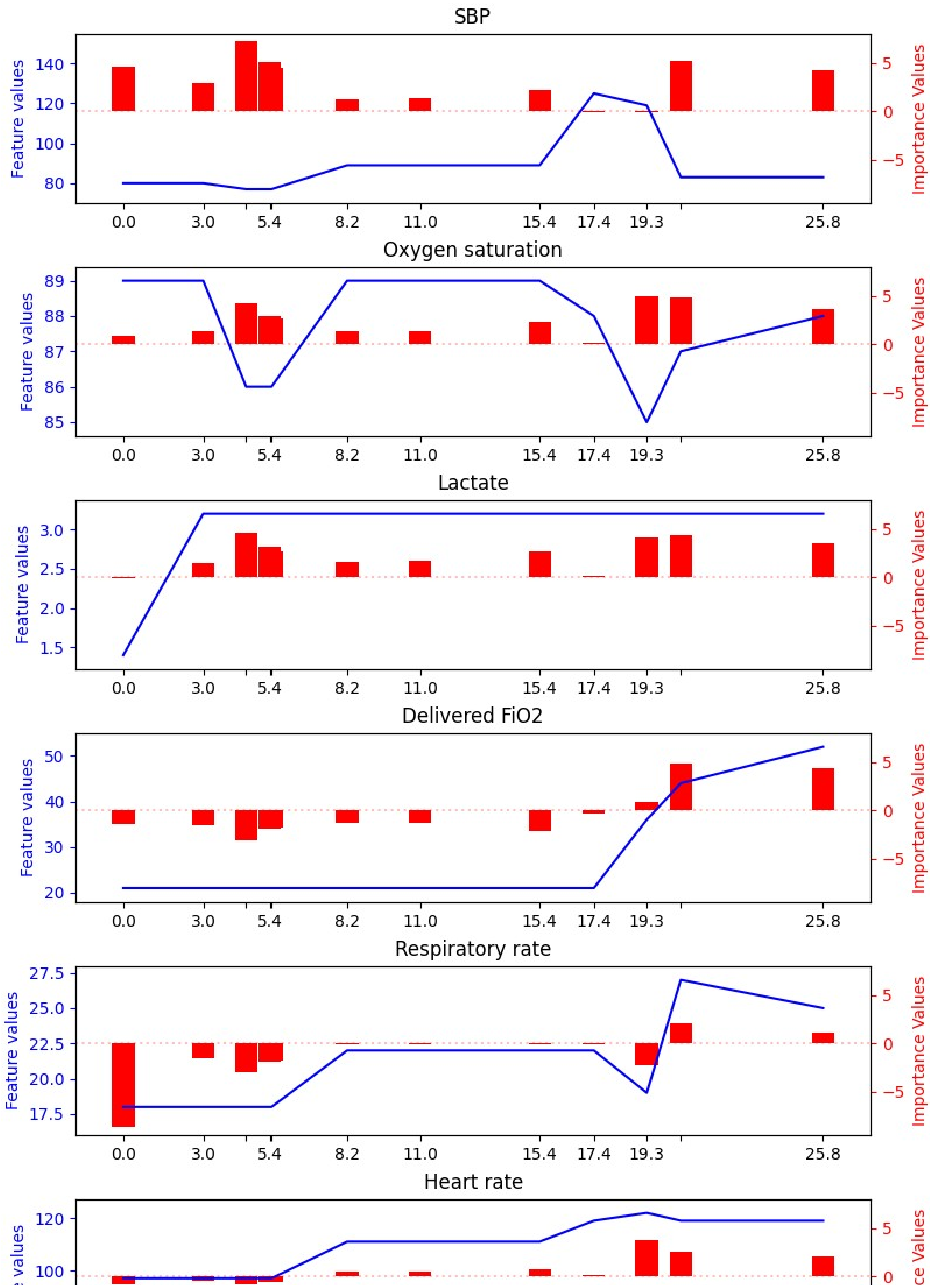
SBP
Feature values
Importance Values
140
120
100
80
5
0
−5
0.0
3.0
5.4
8.2
11.0
15.4
17.4
19.3
25.8
Oxygen saturation
89
88
87
86
85
Lactate
3.0
2.5
2.0
1.5
Delivered FiO2
50
40
30
20
Respiratory rate
27.5
25.0
22.5
20.0
17.5
Heart rate
120
100

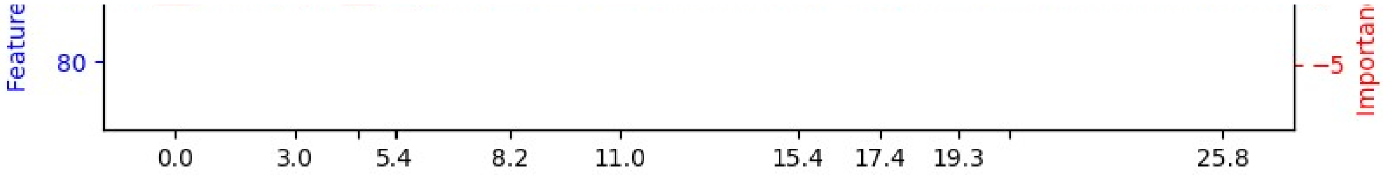


**Before answering the questions below, would it be helpful to see the vital signs, lab results, and description of the patient again?**

- Yes
- No

**Before answering the questions below, would it be helpful to see the description of an attribution explanation again?**

- Yes
- No

**For the features shown below, please rank the features from the most important (that you would use to make a prediction about this patient's likelihood of clinical deterioration in the next 24 hours) to the least important.**

| (One selection allowed per column) | Most Important (1) | Second-most Important (2) | Third-most Important (3) | Fourth-most Important (4) | Fifth-most Important (5) |
|---|---|---|---|---|---|
| **Heart rate** | ○ | ○ | ○ | ○ | ○ |
| **Respiratory rate** | ○ | ○ | ○ | ○ | ○ |
| **SBP** | ○ | ○ | ○ | ○ | ○ |
| **DBP** | ○ | ○ | ○ | ○ | ○ |
| **Oxygen saturation** | ○ | ○ | ○ | ○ | ○ |
| **Temperature (Celsius)** | ○ | ○ | ○ | ○ | ○ |
| **AVPU mental status** | ○ | ○ | ○ | ○ | ○ |
| **Disoriented** | ○ | ○ | ○ | ○ | ○ |
| **Delivered FiO2** | ○ | ○ | ○ | ○ | ○ |
| **Braden Scale - Nutrition** | ○ | ○ | ○ | ○ | ○ |
| **Sum Total of Braden Scale** | ○ | ○ | ○ | ○ | ○ |
| **Lactate** | ○ | ○ | ○ | ○ | ○ |
| **Creatinine** | ○ | ○ | ○ | ○ | ○ |

**Please select one of the options below to indicate your level of agreement with the statement: "I understand the explanation of the machine learning model's prediction for this patient case."**

| I agree strongly | I agree somewhat | I'm neutral about it | I disagree somewhat | I disagree strongly |
|---|---|---|---|---|
| ○ | ○ | ○ | ○ | ○ |

**Please select one of the options below to indicate your level of agreement with the statement: "The explanation increases my trust in the machine learning model's predictions."**

| I agree strongly | I agree somewhat | I'm neutral about it | I disagree somewhat | I disagree strongly |
|---|---|---|---|---|
| ○ | ○ | ○ | ○ | ○ |

**Please select one of the options below to indicate your level of agreement with the statement: "The explanation increased my understanding of how the machine learning model made a prediction for this patient."**

| | I agree strongly | I agree somewhat | I'm neutral about it | I disagree somewhat | I disagree strongly |
|---|---|---|---|---|---|
| | ○ | ○ | ○ | ○ | ○ |

<< Previous Page

Next Page >>

Save & Return Later

Powered by REDCap

### User Preferences for Machine Learning Model Explanations

AAA



### Survey Question 1

**Here we show a rule-based explanation for why the patient has been identified as being at high risk of clinical deterioration.**

**BECAUSE:**

Lactate at time 20.5 > 1.4
Lactate at time 25.8 > 1.4
Delivered FiO2 at time 25.8 > 21.0
Respiratory rate at time 25.8 > 20
Heart rate at time 25.8 > 92.0
Braden scale at time 8.2 <= 19.0

**THEN:**

This patient is expected to be at high risk of clinical deterioration.

**Before answering the questions below, would it be helpful to see the vital signs, lab results, and description of the patient again?**

- Yes
- No

**Before answering the questions below, would it be helpful to see the description of a rule-based explanation again?**

- Yes
- No

**For the features shown below, please rank the features from the most important (that you would use to make a prediction about this patient's likelihood of clinical deterioration in the next 24 hours) to the least important.**

| (One selection allowed per column) | Most Important (1) | Second-most Important (2) | Third-most Important (3) | Fourth-most Important (4) | Fifth-most Important (5) |
|---|---|---|---|---|---|
| **Heart rate** | ○ | ○ | ○ | ○ | ○ |

| | | | | | |
|---|---|---|---|---|---|
| **Respiratory rate** | ○ | ○ | ○ | ○ | ○ |
| **SBP** | ○ | ○ | ○ | ○ | ○ |
| **DBP** | ○ | ○ | ○ | ○ | ○ |
| **Oxygen saturation** | ○ | ○ | ○ | ○ | ○ |
| **Temperature (Celsius)** | ○ | ○ | ○ | ○ | ○ |
| **AVPU mental status** | ○ | ○ | ○ | ○ | ○ |
| **Disoriented** | ○ | ○ | ○ | ○ | ○ |
| **Delivered FiO2** | ○ | ○ | ○ | ○ | ○ |
| **Braden Scale - Nutrition** | ○ | ○ | ○ | ○ | ○ |
| **Sum Total of Braden Scale** | ○ | ○ | ○ | ○ | ○ |
| **Lactate** | ○ | ○ | ○ | ○ | ○ |
| **Creatinine** | ○ | ○ | ○ | ○ | ○ |

**Please select one of the options below to indicate your level of agreement with the statement: "I understand the explanation of the machine learning model's prediction for this patient case."**

| I agree strongly | I agree somewhat | I'm neutral about it | I disagree somewhat | I disagree strongly |
|---|---|---|---|---|
| ○ | ○ | ○ | ○ | ○ |

**Please select one of the options below to indicate your level of agreement with the statement: "The explanation increases my trust in the machine learning model's predictions."**

| I agree strongly | I agree somewhat | I'm neutral about it | I disagree somewhat | I disagree strongly |
|---|---|---|---|---|
| ○ | ○ | ○ | ○ | ○ |

**Please select one of the options below to indicate your level of agreement with the statement: "The explanation increased my understanding of how the machine learning model made a prediction for this patient."**

| I agree strongly | I agree somewhat | I'm neutral about it | I disagree somewhat | I disagree strongly |
|---|---|---|---|---|
| ○ | ○ | ○ | ○ | ○ |

<< Previous Page

Next Page >>

Save & Return Later

Powered by REDCap

**User Preferences for Machine Learning Model Explanations**

A A A



### Survey Question 1

**Here we show a counterfactual explanation for why the patient has been identified as being at high risk of clinical deterioration.**

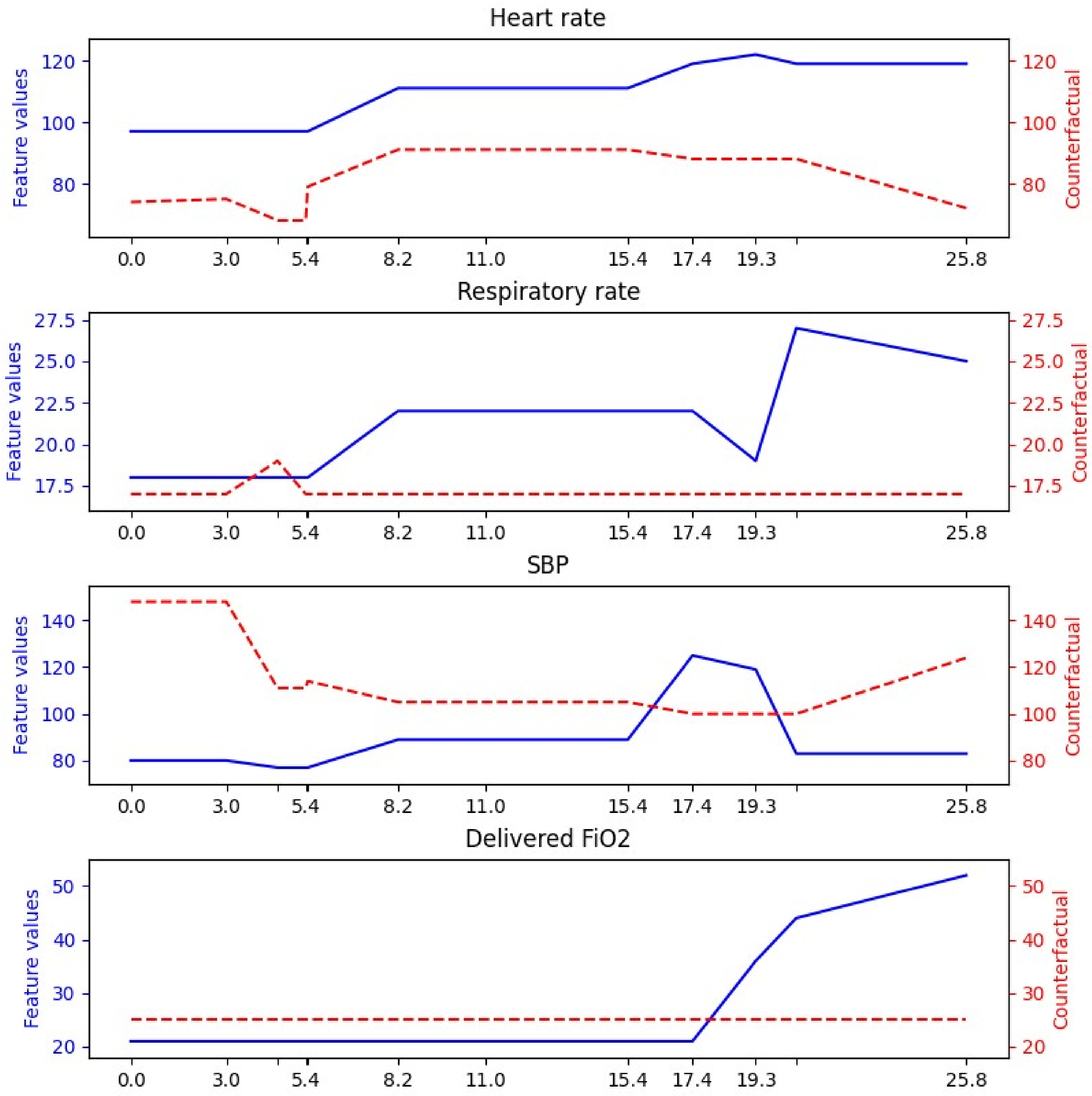


**Before answering the questions below, would it be helpful to see the vital signs, lab results, and description of the patient again?**

- ○ Yes
- ○ No

**Before answering the questions below, would it be helpful to see the description of a counterfactual explanation again?**

- ○ Yes
- ○ No

**For the features shown below, please rank the features from the most important (that you would use to make a prediction about this patient's likelihood of clinical deterioration in the next 24 hours) to the least important.**

| (One selection allowed per column) | Most Important (1) | Second-most Important (2) | Third-most Important (3) | Fourth-most Important (4) | Fifth-most Important (5) |
|---|---|---|---|---|---|
| **Heart rate** | ○ | ○ | ○ | ○ | ○ |
| **Respiratory rate** | ○ | ○ | ○ | ○ | ○ |
| **SBP** | ○ | ○ | ○ | ○ | ○ |
| **DBP** | ○ | ○ | ○ | ○ | ○ |
| **Oxygen saturation** | ○ | ○ | ○ | ○ | ○ |
| **Temperature (Celsius)** | ○ | ○ | ○ | ○ | ○ |
| **AVPU mental status** | ○ | ○ | ○ | ○ | ○ |
| **Disoriented** | ○ | ○ | ○ | ○ | ○ |
| **Delivered FiO2** | ○ | ○ | ○ | ○ | ○ |
| **Braden Scale - Nutrition** | ○ | ○ | ○ | ○ | ○ |
| **Sum Total of Braden Scale** | ○ | ○ | ○ | ○ | ○ |
| **Lactate** | ○ | ○ | ○ | ○ | ○ |
| **Creatinine** | ○ | ○ | ○ | ○ | ○ |

**Please select one of the options below to indicate your level of agreement with the statement: "I understand the explanation of the machine learning model's prediction for this patient case."**

| I agree strongly | I agree somewhat | I'm neutral about it | I disagree somewhat | I disagree strongly |
|---|---|---|---|---|
| ○ | ○ | ○ | ○ | ○ |

**Please select one of the options below to indicate your level of agreement with the statement: "The explanation increases my trust in the machine learning model's predictions."**

| I agree strongly | I agree somewhat | I'm neutral about it | I disagree somewhat | I disagree strongly |
|---|---|---|---|---|
| ○ | ○ | ○ | ○ | ○ |

**Please select one of the options below to indicate your level of agreement with the statement: "The explanation increased my understanding of how the machine learning model made a prediction for this patient."**

| I agree strongly | I agree somewhat | I'm neutral about it | I disagree somewhat | I disagree strongly |
|---|---|---|---|---|
| ○ | ○ | ○ | ○ | ○ |

<< Previous Page

Next Page >>

Save & Return Later

Powered by REDCap

#### User Preferences for Machine Learning Model Explanations

AAA



### Survey Question 2

**You have received an alert that a patient has been identified as being at high risk of clinical deterioration. For this patie the vital signs, lab results, and a description of the patient are displayed below.**

**Clinical Notes:**

The patient is a 74-year-old male with a BMI of 39.8

**The features given to the machine learning model are shown below.**

| Hours Since Admit | 6.2 | 6.5 | 7.2 | 8 | 12.4 | 32 | 34.9 | 40.6 | 40.9 | 41 | 4 |
|---|---|---|---|---|---|---|---|---|---|---|---|
| **Vitals** | | | | | | | | | | | |
| Heart rate | - | 130 | 71 | 69 | 92 | 70 | - | 104 | - | 130 | 1 |
| Respiratory rate | 24 | 24 | 22 | 22 | - | 21 | - | - | 40 | 40 | [illegible] |
| SBP | - | 130 | - | 137 | 117 | 142 | - | 136 | - | 140 | [illegible] |
| DBP | - | 74 | - | 67 | 75 | 83 | - | 78 | - | 76 | [illegible] |
| Oxygen saturation | - | 95 | 98 | 96 | 95 | 93 | 94 | 89 | - | 92 | [illegible] |
| Temperature (Celsius) | - | 37.8 | - | 38.6 | 36.9 | 38.2 | - | 39.8 | - | 39.7 | 3 |
| AVPU mental status | Alert | - | - | - | - | - | - | - | Responds to Voice | - | [illegible] |
| Disoriented | - | - | - | - | - | - | - | - | Yes | - | [illegible] |
| Delivered FiO2 | - | 31 | 31 | - | - | 31 | 31 | 31 | - | 31 | [illegible] |
| Braden Scale - Nutrition | 2 | - | - | - | - | - | - | - | 2 | - | [illegible] |
| Sum Total of Braden Scale | 14 | - | - | - | - | - | - | - | 14 | - | [illegible] |
| **Labs** | | | | | | | | | | | |
| Lactate | - | - | - | - | - | - | - | - | - | - | [illegible] |
| Creatinine | - | - | - | - | - | - | - | - | - | - | [illegible] |

**For the features shown, please rank what you think are the five most important features (that you would use to make a prediction about this patient's likelihood of clinical deterioration in the next 24 hours).**

| (One selection allowed per column) | Most Important (1) | Second-most Important (2) | Third-most Important (3) | Fourth-most Important (4) | Fifth-mo Importan |
|---|---|---|---|---|---|
| Heart rate | ○ | ○ | ○ | ○ | ○ |
| Respiratory rate | ○ | ○ | ○ | ○ | ○ |
| SBP | ○ | ○ | ○ | ○ | ○ |
| DBP | ○ | ○ | ○ | ○ | ○ |
| Oxygen saturation | ○ | ○ | ○ | ○ | ○ |
| Temperature (Celsius) | ○ | ○ | ○ | ○ | ○ |

| | | | | | |
|---|---|---|---|---|---|
| **AVPU mental status** | ○ | ○ | ○ | ○ | ○ |
| **Disoriented** | ○ | ○ | ○ | ○ | ○ |
| **Delivered FiO2** | ○ | ○ | ○ | ○ | ○ |
| **Braden Scale - Nutrition** | ○ | ○ | ○ | ○ | ○ |
| **Sum Total of Braden Scale** | ○ | ○ | ○ | ○ | ○ |
| **Lactate** | ○ | ○ | ○ | ○ | ○ |
| **Creatinine** | ○ | ○ | ○ | ○ | ○ |

**<< Previous Page**

**Next Page >>**

**Save & Return Later**

Powered by REDCap

### User Preferences for Machine Learning Model Explanations

A A A

Page 9 of 26

### Survey Question 2

**Here we show a rule-based explanation for why the patient has been identified as being at high risk of clinical deterioration.**

**BECAUSE:**

Respiratory rate at time 41.6 > 20.0

Delivered FiO2 at time 41.6 > 21.0

**THEN:**

This patient is expected to be at high risk of clinical deterioration.

**Before answering the questions below, would it be helpful to see the vital signs, lab results, and description of the patient again?**

- Yes
- No

**Before answering the questions below, would it be helpful to see the description of a rule-based explanation again?**

- Yes
- No

**For the features shown below, please rank the features from the most important (that you would use to make a prediction about this patient's likelihood of clinical deterioration in the next 24 hours) to the least important.**

| (One selection allowed per column) | Most Important (1) | Second-most Important (2) | Third-most Important (3) | Fourth-most Important (4) | Fifth-most Important (5) |
|---|---|---|---|---|---|
| **Heart rate** | ○ | ○ | ○ | ○ | ○ |
| **Respiratory rate** | ○ | ○ | ○ | ○ | ○ |
| **SBP** | ○ | ○ | ○ | ○ | ○ |
| **DBP** | ○ | ○ | ○ | ○ | ○ |

| | | | | | |
|---|---|---|---|---|---|
| **Oxygen saturation** | ○ | ○ | ○ | ○ | ○ |
| **Temperature (Celsius)** | ○ | ○ | ○ | ○ | ○ |
| **AVPU mental status** | ○ | ○ | ○ | ○ | ○ |
| **Disoriented** | ○ | ○ | ○ | ○ | ○ |
| **Delivered FiO2** | ○ | ○ | ○ | ○ | ○ |
| **Braden Scale - Nutrition** | ○ | ○ | ○ | ○ | ○ |
| **Sum Total of Braden Scale** | ○ | ○ | ○ | ○ | ○ |
| **Lactate** | ○ | ○ | ○ | ○ | ○ |
| **Creatinine** | ○ | ○ | ○ | ○ | ○ |

**Please select one of the options below to indicate your level of agreement with the statement: "I understand the explanation of the machine learning model's prediction for this patient case."**

| | I agree strongly | I agree somewhat | I'm neutral about it | I disagree somewhat | I disagree strongly |
|---|---|---|---|---|---|
| | ○ | ○ | ○ | ○ | ○ |

**Please select one of the options below to indicate your level of agreement with the statement: "The explanation increases my trust in the machine learning model's predictions."**

| | I agree strongly | I agree somewhat | I'm neutral about it | I disagree somewhat | I disagree strongly |
|---|---|---|---|---|---|
| | ○ | ○ | ○ | ○ | ○ |

**Please select one of the options below to indicate your level of agreement with the statement: "The explanation increased my understanding of how the machine learning model made a prediction for this patient."**

| | I agree strongly | I agree somewhat | I'm neutral about it | I disagree somewhat | I disagree strongly |
|---|---|---|---|---|---|
| | ○ | ○ | ○ | ○ | ○ |

<< Previous Page

Next Page >>

Save & Return Later

Powered by REDCap

**User Preferences for Machine Learning Model Explanations**

A A A

Page 10 of 26

### Survey Question 2

**Here we show an attribution explanation for why the patient has been identified as being at high risk of clinical deterioration.**

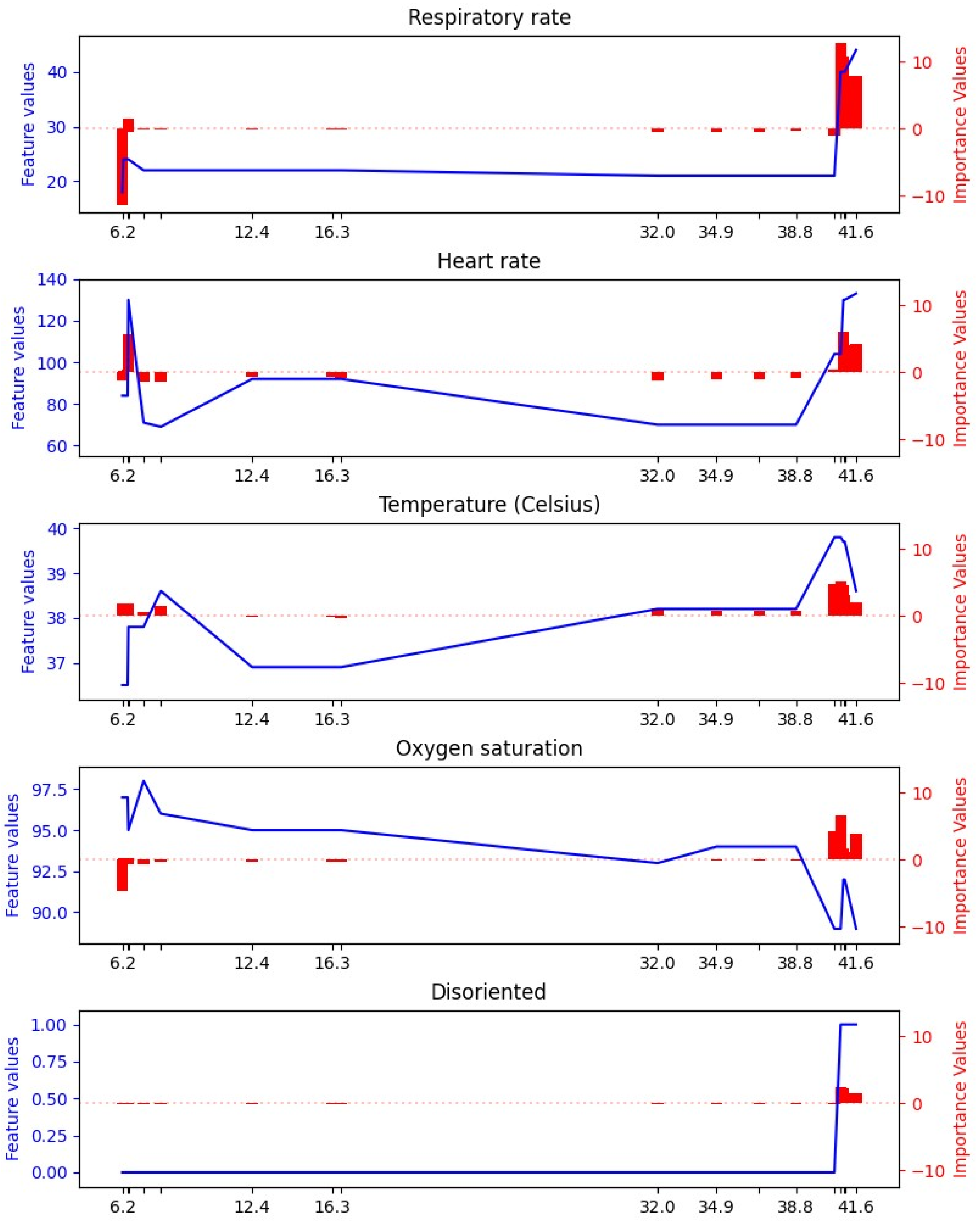
Respiratory rate
Heart rate
Temperature (Celsius)
Oxygen saturation
Disoriented
Feature values
Importance Values
6.2
12.4
16.3
32.0
34.9
38.8
41.6

**Before answering the questions below, would it be helpful to see the vital signs, lab results, and description of the patient again?**

- ○ Yes
- ○ No

**Before answering the questions below, would it be helpful to see the description of an attribution explanation again?**

- ○ Yes
- ○ No

**For the features shown below, please rank the features from the most important (that you would use to make a prediction about this patient's likelihood of clinical deterioration in the next 24 hours) to the least important.**

| (One selection allowed per column) | Most Important (1) | Second-most Important (2) | Third-most Important (3) | Fourth-most Important (4) | Fifth-most Important (5) |
|---|---|---|---|---|---|
| **Heart rate** | ○ | ○ | ○ | ○ | ○ |
| **Respiratory rate** | ○ | ○ | ○ | ○ | ○ |
| **SBP** | ○ | ○ | ○ | ○ | ○ |
| **DBP** | ○ | ○ | ○ | ○ | ○ |
| **Oxygen saturation** | ○ | ○ | ○ | ○ | ○ |
| **Temperature (Celsius)** | ○ | ○ | ○ | ○ | ○ |
| **AVPU mental status** | ○ | ○ | ○ | ○ | ○ |
| **Disoriented** | ○ | ○ | ○ | ○ | ○ |
| **Delivered FiO2** | ○ | ○ | ○ | ○ | ○ |
| **Braden Scale - Nutrition** | ○ | ○ | ○ | ○ | ○ |
| **Sum Total of Braden Scale** | ○ | ○ | ○ | ○ | ○ |
| **Lactate** | ○ | ○ | ○ | ○ | ○ |
| **Creatinine** | ○ | ○ | ○ | ○ | ○ |

**Please select one of the options below to indicate your level of agreement with the statement: "I understand the explanation of the machine learning model's prediction for this patient case."**

| I agree strongly | I agree somewhat | I'm neutral about it | I disagree somewhat | I disagree strongly |
|---|---|---|---|---|
| ○ | ○ | ○ | ○ | ○ |

**Please select one of the options below to indicate your level of agreement with the statement: "The explanation increases my trust in the machine learning model's predictions."**

| I agree strongly | I agree somewhat | I'm neutral about it | I disagree somewhat | I disagree strongly |
|---|---|---|---|---|
| ○ | ○ | ○ | ○ | ○ |

**Please select one of the options below to indicate your level of agreement with the statement: "The explanation increased my understanding of how the machine learning model made a prediction for this patient."**

| | I agree strongly | I agree somewhat | I'm neutral about it | I disagree somewhat | I disagree strongly |
|---|---|---|---|---|---|
| | ○ | ○ | ○ | ○ | ○ |

<< Previous Page

Next Page >>

Save & Return Later

Powered by REDCap

**User Preferences for Machine Learning Model Explanations**

AAA

Page 11 of 26

### Survey Question 2

**Here we show a counterfactual explanation for why the patient has been identified as being at high risk of clinical deterioration.**

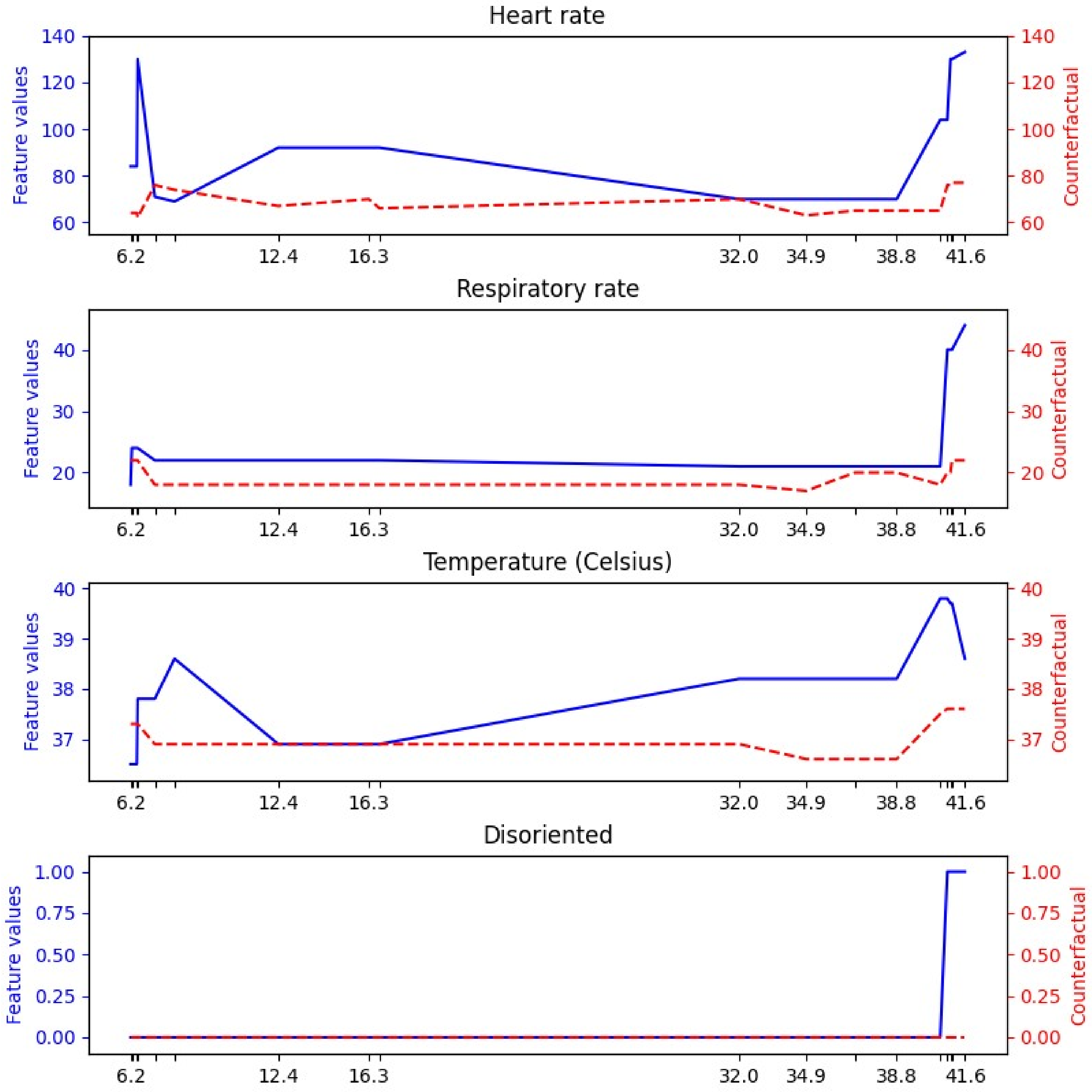


**Before answering the questions below, would it be helpful to see the vital signs, lab results, and description of the patient again?**

- ○ Yes
- ○ No

**Before answering the questions below, would it be helpful to see the description of a counterfactual explanation again?**

- ○ Yes
- ○ No

**For the features shown below, please rank the features from the most important (that you would use to make a prediction about this patient's likelihood of clinical deterioration in the next 24 hours) to the least important.**

| (One selection allowed per column) | Most Important (1) | Second-most Important (2) | Third-most Important (3) | Fourth-most Important (4) | Fifth-most Important (5) |
|---|---|---|---|---|---|
| **Heart rate** | ○ | ○ | ○ | ○ | ○ |
| **Respiratory rate** | ○ | ○ | ○ | ○ | ○ |
| **SBP** | ○ | ○ | ○ | ○ | ○ |
| **DBP** | ○ | ○ | ○ | ○ | ○ |
| **Oxygen saturation** | ○ | ○ | ○ | ○ | ○ |
| **Temperature (Celsius)** | ○ | ○ | ○ | ○ | ○ |
| **AVPU mental status** | ○ | ○ | ○ | ○ | ○ |
| **Disoriented** | ○ | ○ | ○ | ○ | ○ |
| **Delivered FiO2** | ○ | ○ | ○ | ○ | ○ |
| **Braden Scale - Nutrition** | ○ | ○ | ○ | ○ | ○ |
| **Sum Total of Braden Scale** | ○ | ○ | ○ | ○ | ○ |
| **Lactate** | ○ | ○ | ○ | ○ | ○ |
| **Creatinine** | ○ | ○ | ○ | ○ | ○ |

**Please select one of the options below to indicate your level of agreement with the statement: "I understand the explanation of the machine learning model's prediction for this patient case."**

| I agree strongly | I agree somewhat | I'm neutral about it | I disagree somewhat | I disagree strongly |
|---|---|---|---|---|
| ○ | ○ | ○ | ○ | ○ |

**Please select one of the options below to indicate your level of agreement with the statement: "The explanation increases my trust in the machine learning model's predictions."**

| I agree strongly | I agree somewhat | I'm neutral about it | I disagree somewhat | I disagree strongly |
|---|---|---|---|---|
| ○ | ○ | ○ | ○ | ○ |

**Please select one of the options below to indicate your level of agreement with the statement: "The explanation increased my understanding of how the machine learning model made a prediction for this patient."**

| I agree strongly | I agree somewhat | I'm neutral about it | I disagree somewhat | I disagree strongly |
|---|---|---|---|---|
| ○ | ○ | ○ | ○ | ○ |

<< Previous Page

Next Page >>

Save & Return Later

Powered by REDCap

#### User Preferences for Machine Learning Model Explanations

AAA

Page 12 of 26

### Survey Question 3

**You have received an alert that a patient has been identified as being at high risk of clinical deterioration. For this patient, the vital signs, lab results, a[illegible] are displayed below.**

**Clinical Notes:**

The patient is a 67-year-old male with a BMI of 43.8

**The features given to the machine learning model are shown below.**

| Hours Since Admit | 0 | 0.3 | 1.6 | 5.3 | 13.3 | 14.3 | 17.3 | 17.8 | 24.3 | 27.6 | 29.3 | 33.3 | [illegible] |
|---|---|---|---|---|---|---|---|---|---|---|---|---|---|
| **Vitals** | | | | | | | | | | | | | |
| Heart rate | 106 | - | - | 106 | - | 103 | - | 103 | - | 109 | 115 | 113 | [illegible] |
| Respiratory rate | - | 22 | - | 22 | - | 22 | - | - | - | - | - | 20 | [illegible] |
| SBP | 108 | - | - | 80 | - | 111 | - | 107 | - | 79 | - | 97 | [illegible] |
| DBP | 45 | - | - | 40 | - | 44 | - | 48 | - | 42 | - | 56 | [illegible] |
| Oxygen saturation | 91 | - | - | 89 | - | 92 | - | 95 | - | - | - | 95 | [illegible] |
| Temperature (Celsius) | 37.2 | - | - | 37.2 | - | 37.3 | - | - | - | - | - | 37.2 | [illegible] |
| AVPU mental status | - | Responds to Voice | - | Responds to Voice | Responds to Voice | - | Responds to Voice | - | Responds to Voice | - | Responds to Voice | Responds to Pain | [illegible] |
| Disoriented | - | No | - | No | No | - | No | - | No | - | No | No | [illegible] |
| Delivered FiO2 | 32 | - | - | 32 | - | 32 | - | 32 | - | - | - | 32 | [illegible] |
| Braden Scale - Nutrition | - | 2 | - | - | - | - | - | - | - | - | - | 2 | [illegible] |
| Sum Total of Braden Scale | - | 16 | - | - | - | - | - | - | - | - | - | 17 | [illegible] |
| **Labs** | | | | | | | | | | | | | |
| Lactate | - | - | - | - | - | - | - | - | - | - | - | - | [illegible] |
| Creatinine | - | - | 2.5 | - | - | - | - | - | - | - | - | - | [illegible] |

**For the features shown, please rank what you think are the five most important features (that you would use to make a prediction about this patient'[illegible] deterioration in the next 24 hours).**

| (One selection allowed per column) | Most Important (1) | Second-most Important (2) | Third-most Important (3) | Fourth-most Im[illegible] |
|---|---|---|---|---|
| Heart rate | ○ | ○ | ○ | ○ |
| Respiratory rate | ○ | ○ | ○ | ○ |
| SBP | ○ | ○ | ○ | ○ |
| DBP | ○ | ○ | ○ | ○ |
| Oxygen saturation | ○ | ○ | ○ | ○ |
| Temperature (Celsius) | ○ | ○ | ○ | ○ |
| AVPU mental status | ○ | ○ | ○ | ○ |
| Disoriented | ○ | ○ | ○ | ○ |
| Delivered FiO2 | ○ | ○ | ○ | ○ |
| Braden Scale - Nutrition | ○ | ○ | ○ | ○ |
| Sum Total of Braden Scale | ○ | ○ | ○ | ○ |
| Lactate | ○ | ○ | ○ | ○ |
| Creatinine | ○ | ○ | ○ | ○ |

<< Previous Page

Next Page >>

Save & Return Later

Powered by REDCap

**User Preferences for Machine Learning Model Explanations**

AAA

**Page 13 of 26**

### Survey Question 3

**Here we show an attribution explanation for why the patient has been identified as being at high risk of clinical deterioration.**

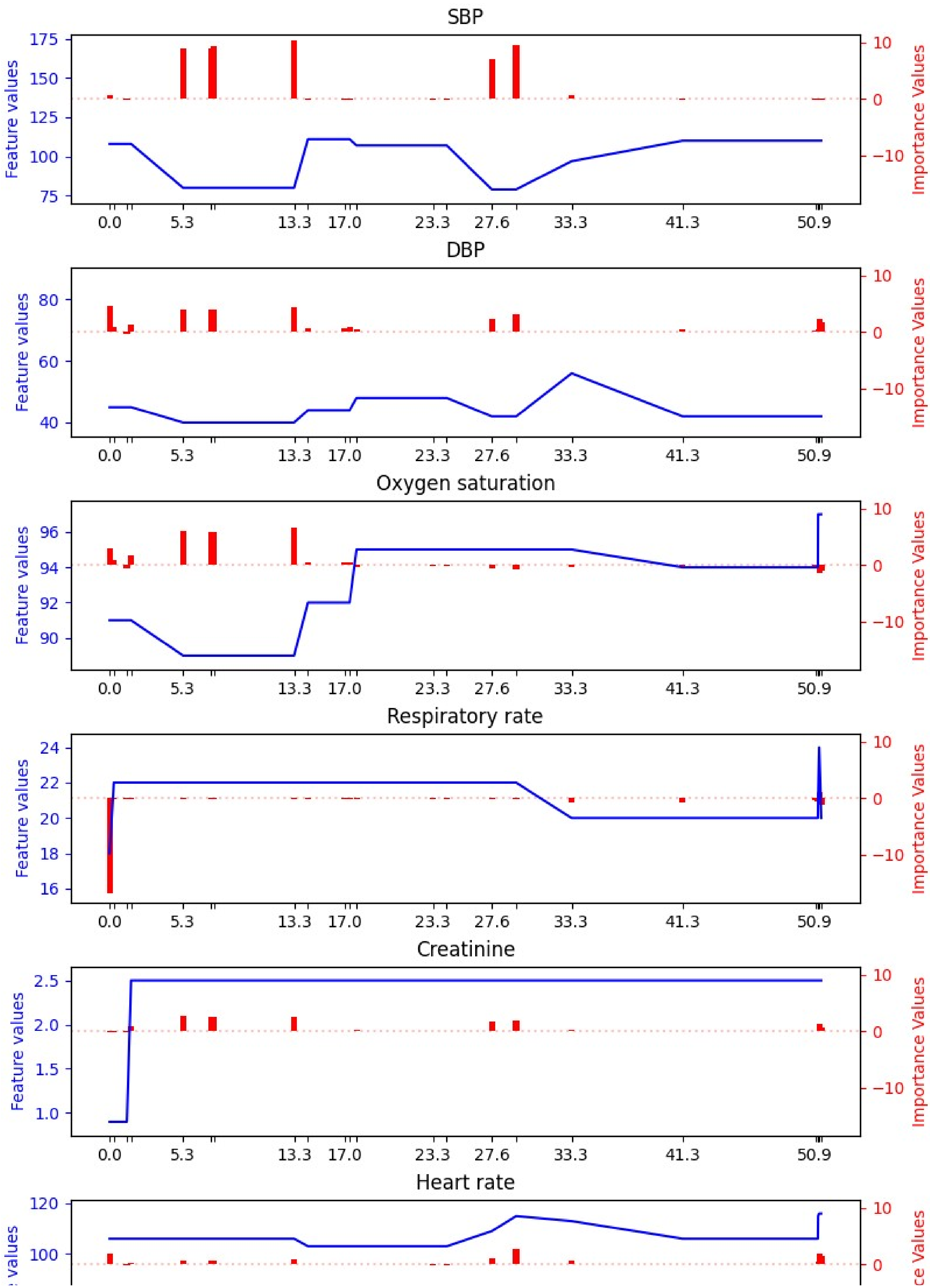
SBP
Feature values
Importance Values
175
150
125
100
75
10
0
−10
0.0
5.3
13.3
17.0
23.3
27.6
33.3
41.3
50.9
DBP
Feature values
Importance Values
80
60
40
Oxygen saturation
Feature values
Importance Values
96
94
92
90
Respiratory rate
Feature values
Importance Values
24
22
20
18
16
Creatinine
Feature values
Importance Values
2.5
2.0
1.5
1.0
Heart rate
120
100

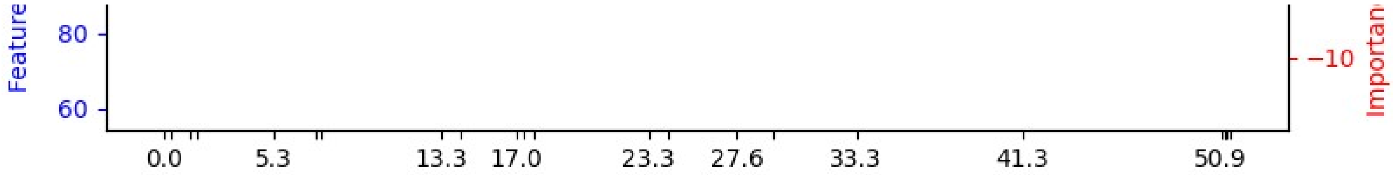


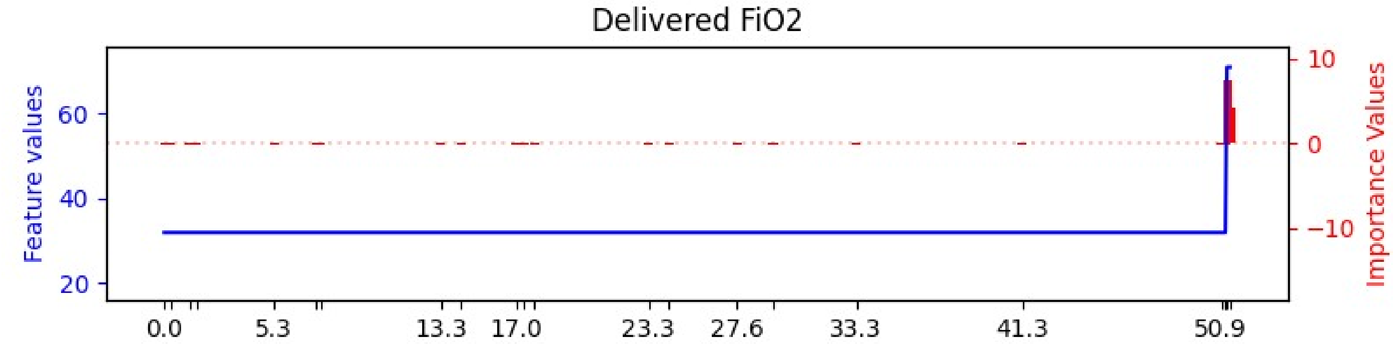


**Before answering the questions below, would it be helpful to see the vital signs, lab results, and description of the patient again?**

- Yes
- No

**Before answering the questions below, would it be helpful to see the description of an attribution explanation again?**

- Yes
- No

**For the features shown below, please rank the features from the most important (that you would use to make a prediction about this patient's likelihood of clinical deterioration in the next 24 hours) to the least important.**

| (One selection allowed per column) | Most Important (1) | Second-most Important (2) | Third-most Important (3) | Fourth-most Important (4) | Fifth-most Important (5) |
|---|---|---|---|---|---|
| **Heart rate** | | | | | |
| **Respiratory rate** | | | | | |
| **SBP** | | | | | |
| **DBP** | | | | | |
| **Oxygen saturation** | | | | | |
| **Temperature (Celsius)** | | | | | |
| **AVPU mental status** | | | | | |
| **Disoriented** | | | | | |
| **Delivered FiO2** | | | | | |
| **Braden Scale - Nutrition** | | | | | |
| **Sum Total of Braden Scale** | | | | | |
| **Lactate** | | | | | |
| **Creatinine** | | | | | |

**Please select one of the options below to indicate your level of agreement with the statement: "I understand the explanation of the machine learning model's prediction for this patient case."**

| | I agree strongly | I agree somewhat | I'm neutral about it | I disagree somewhat | I disagree strongly |
|---|---|---|---|---|---|
| | ○ | ○ | ○ | ○ | ○ |

**Please select one of the options below to indicate your level of agreement with the statement: "The explanation increases my trust in the machine learning model's predictions."**

| | I agree strongly | I agree somewhat | I'm neutral about it | I disagree somewhat | I disagree strongly |
|---|---|---|---|---|---|
| | ○ | ○ | ○ | ○ | ○ |

**Please select one of the options below to indicate your level of agreement with the statement: "The explanation increased my understanding of how the machine learning model made a prediction for this patient."**

| | I agree strongly | I agree somewhat | I'm neutral about it | I disagree somewhat | I disagree strongly |
|---|---|---|---|---|---|
| | ○ | ○ | ○ | ○ | ○ |

<< Previous Page

Next Page >>

Save & Return Later

Powered by REDCap

### User Preferences for Machine Learning Model Explanations

A A A

**Page 14 of 26**

### Survey Question 3

**Here we show a rule-based explanation for why the patient has been identified as being at high risk of clinical deterioration.**

**BECAUSE:**

Delivered FiO2 at time 51.0 > 21.0

Heart rate at time 51.0 > 92.0

Braden Scale - Nutrition at time 27.5 = 2

**THEN:**

This patient is expected to be at high risk of clinical deterioration.

**Before answering the questions below, would it be helpful to see the vital signs, lab results, and description of the patient again?**

- Yes
- No

**Before answering the questions below, would it be helpful to see the description of rule-based explanation again?**

- Yes
- No

**For the features shown below, please rank the features from the most important (that you would use to make a prediction about this patient's likelihood of clinical deterioration in the next 24 hours) to the least important.**

| (One selection allowed per column) | Most Important (1) | Second-most Important (2) | Third-most Important (3) | Fourth-most Important (4) | Fifth-most Important (5) |
|---|---|---|---|---|---|
| **Heart rate** | ○ | ○ | ○ | ○ | ○ |
| **Respiratory rate** | ○ | ○ | ○ | ○ | ○ |
| **SBP** | ○ | ○ | ○ | ○ | ○ |

| | | | | | |
|---|---|---|---|---|---|
| **DBP** | ○ | ○ | ○ | ○ | ○ |
| **Oxygen saturation** | ○ | ○ | ○ | ○ | ○ |
| **Temperature (Celsius)** | ○ | ○ | ○ | ○ | ○ |
| **AVPU mental status** | ○ | ○ | ○ | ○ | ○ |
| **Disoriented** | ○ | ○ | ○ | ○ | ○ |
| **Delivered FiO2** | ○ | ○ | ○ | ○ | ○ |
| **Braden Scale - Nutrition** | ○ | ○ | ○ | ○ | ○ |
| **Sum Total of Braden Scale** | ○ | ○ | ○ | ○ | ○ |
| **Lactate** | ○ | ○ | ○ | ○ | ○ |
| **Creatinine** | ○ | ○ | ○ | ○ | ○ |

**Please select one of the options below to indicate your level of agreement with the statement: "I understand the explanation of the machine learning model's prediction for this patient case."**

| I agree strongly | I agree somewhat | I'm neutral about it | I disagree somewhat | I disagree strongly |
|---|---|---|---|---|
| ○ | ○ | ○ | ○ | ○ |

**Please select one of the options below to indicate your level of agreement with the statement: "The explanation increases my trust in the machine learning model's predictions."**

| I agree strongly | I agree somewhat | I'm neutral about it | I disagree somewhat | I disagree strongly |
|---|---|---|---|---|
| ○ | ○ | ○ | ○ | ○ |

**Please select one of the options below to indicate your level of agreement with the statement: "The explanation increased my understanding of how the machine learning model made a prediction for this patient."**

| I agree strongly | I agree somewhat | I'm neutral about it | I disagree somewhat | I disagree strongly |
|---|---|---|---|---|
| ○ | ○ | ○ | ○ | ○ |

<< Previous Page

Next Page >>

Save & Return Later

Powered by REDCap

#### User Preferences for Machine Learning Model Explanations

AAA



### Survey Question 3

**Here we show a counterfactual explanation for why the patient has been identified as being at high risk of clinical deterioration.**

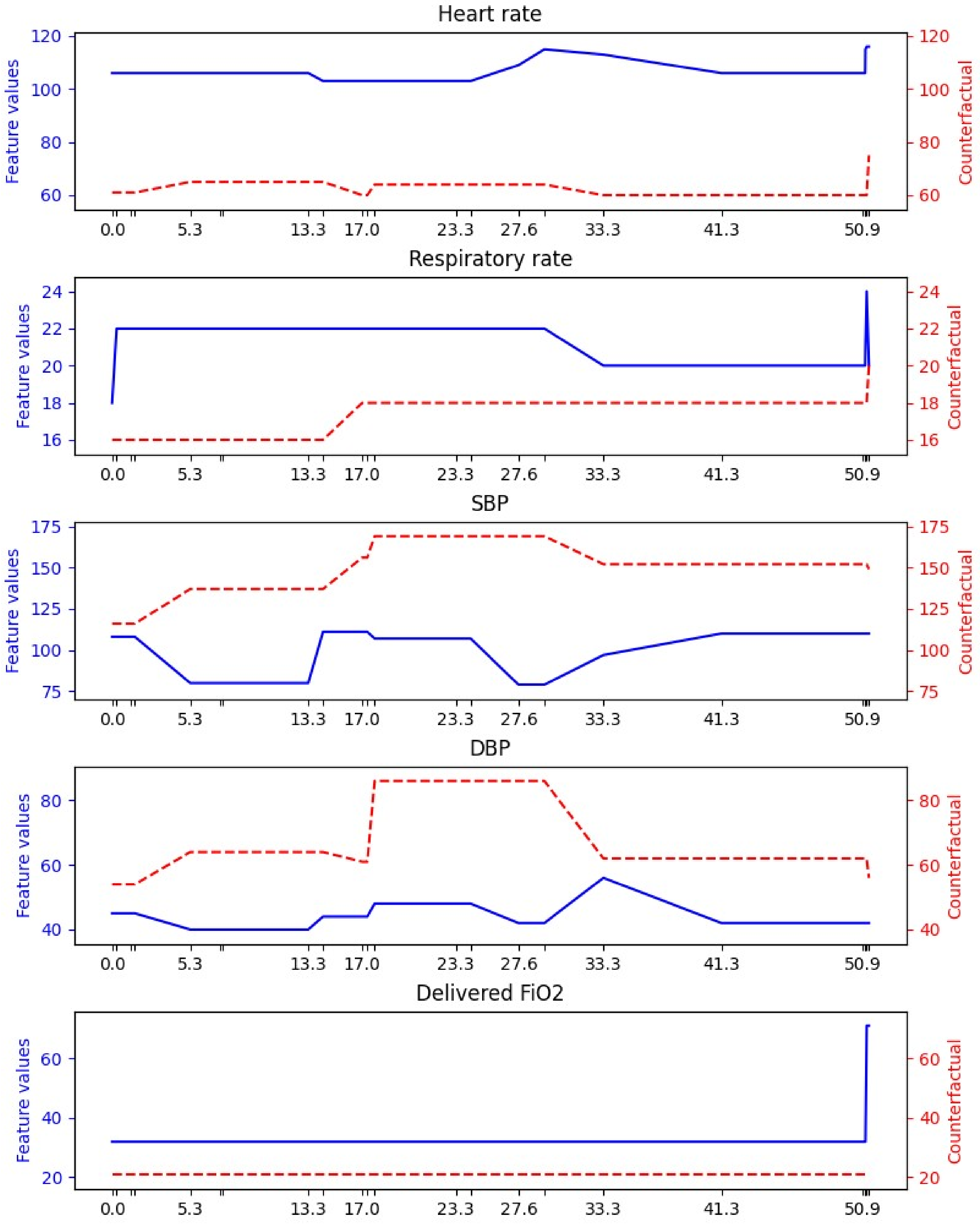
Heart rate
Feature values
Counterfactual
Respiratory rate
Feature values
Counterfactual
SBP
Feature values
Counterfactual
DBP
Feature values
Counterfactual
Delivered FiO2
Feature values
Counterfactual
0.0
5.3
13.3
17.0
23.3
27.6
33.3
41.3
50.9

**Before answering the questions below, would it be helpful to see the vital signs, lab results, and description of the patient again?**

- ○ Yes
- ○ No

**Before answering the questions below, would it be helpful to see the description of a counterfactual explanation again?**

- ○ Yes
- ○ No

**For the features shown below, please rank the features from the most important (that you would use to make a prediction about this patient's likelihood of clinical deterioration in the next 24 hours) to the least important.**

| (One selection allowed per column) | Most Important (1) | Second-most Important (2) | Third-most Important (3) | Fourth-most Important (4) | Fifth-most Important (5) |
|---|---|---|---|---|---|
| **Heart rate** | ○ | ○ | ○ | ○ | ○ |
| **Respiratory rate** | ○ | ○ | ○ | ○ | ○ |
| **SBP** | ○ | ○ | ○ | ○ | ○ |
| **DBP** | ○ | ○ | ○ | ○ | ○ |
| **Oxygen saturation** | ○ | ○ | ○ | ○ | ○ |
| **Temperature (Celsius)** | ○ | ○ | ○ | ○ | ○ |
| **AVPU mental status** | ○ | ○ | ○ | ○ | ○ |
| **Disoriented** | ○ | ○ | ○ | ○ | ○ |
| **Delivered FiO2** | ○ | ○ | ○ | ○ | ○ |
| **Braden Scale - Nutrition** | ○ | ○ | ○ | ○ | ○ |
| **Sum Total of Braden Scale** | ○ | ○ | ○ | ○ | ○ |
| **Lactate** | ○ | ○ | ○ | ○ | ○ |
| **Creatinine** | ○ | ○ | ○ | ○ | ○ |

**Please select one of the options below to indicate your level of agreement with the statement: "I understand the explanation of the machine learning model's prediction for this patient case."**

| I agree strongly | I agree somewhat | I'm neutral about it | I disagree somewhat | I disagree strongly |
|---|---|---|---|---|
| ○ | ○ | ○ | ○ | ○ |

**Please select one of the options below to indicate your level of agreement with the statement: "The explanation increases my trust in the machine learning model's predictions."**

| I agree strongly | I agree somewhat | I'm neutral about it | I disagree somewhat | I disagree strongly |
|---|---|---|---|---|
| ○ | ○ | ○ | ○ | ○ |

**Please select one of the options below to indicate your level of agreement with the statement: "The explanation increased my understanding of how the machine learning model made a prediction for this patient."**

| | I agree strongly | I agree somewhat | I'm neutral about it | I disagree somewhat | I disagree strongly |
| --- | --- | --- | --- | --- | --- |
| | ○ | ○ | ○ | ○ | ○ |

<< Previous Page

Next Page >>

Save & Return Later

Powered by REDCap

### User Preferences for Machine Learning Model Explanations

A A A

**Page 16 of 26**

### Survey Question 4

**You have received an alert that a patient has been identified as being at high risk of clinical deterioration. For this patient, the vital signs, lab results, and a description of the patient are displayed below.**

**Clinical Notes:**

The patient is a 73-year-old male with a BMI of 27.1

**The features given to the machine learning model are shown below.**

| Hours Since Admit | 11.5 | 13.7 | 15.1 | 16 | 26.5 | 31 |
|---|---|---|---|---|---|---|
| **Vitals** | | | | | | |
| Heart rate | - | - | 112 | - | 97 | 107 |
| Respiratory rate | - | 28 | 28 | 28 | 28 | 40 |
| SBP | - | - | 150 | - | - | - |
| DBP | - | - | 79 | - | - | - |
| Oxygen saturation | - | - | 87 | - | 90 | 86 |
| Temperature (Celsius) | - | - | 38.4 | - | - | - |
| AVPU mental status | Responds to Voice | - | - | - | - | - |
| Disoriented | No | - | - | - | - | - |
| Delivered FiO2 | - | 45 | 45 | 49 | 49 | 82 |
| Braden Scale - Nutrition | 2 | - | - | - | - | - |
| Sum Total of Braden Scale | 16 | - | - | - | - | - |
| **Labs** | | | | | | |
| Lactate | - | - | - | - | - | - |
| Creatinine | - | - | - | - | - | - |

**For the features shown, please rank what you think are the five most important features (that you would use to make a prediction about this patient's likelihood of clinical deterioration in the next 24 hours).**

| (One selection allowed per column) | Most Important (1) | Second-most Important (2) | Third-most Important (3) | Fourth-most Important (4) | Fifth-most Important (5) |
|---|---|---|---|---|---|
| **Heart rate** | ○ | ○ | ○ | ○ | ○ |
| **Respiratory rate** | ○ | ○ | ○ | ○ | ○ |
| **SBP** | ○ | ○ | ○ | ○ | ○ |
| **DBP** | ○ | ○ | ○ | ○ | ○ |
| **Oxygen saturation** | ○ | ○ | ○ | ○ | ○ |
| **Temperature (Celsius)** | ○ | ○ | ○ | ○ | ○ |
| **AVPU mental status** | ○ | ○ | ○ | ○ | ○ |
| **Disoriented** | ○ | ○ | ○ | ○ | ○ |
| **Delivered FiO2** | ○ | ○ | ○ | ○ | ○ |
| **Braden Scale - Nutrition** | ○ | ○ | ○ | ○ | ○ |
| **Sum Total of Braden Scale** | ○ | ○ | ○ | ○ | ○ |
| **Lactate** | ○ | ○ | ○ | ○ | ○ |
| **Creatinine** | ○ | ○ | ○ | ○ | ○ |

<< Previous Page

Next Page >>

Save & Return Later

Powered by REDCap

#### User Preferences for Machine Learning Model Explanations

AAA



### Survey Question 4

**Here we show an attribution explanation for why the patient has been identified as being at high risk of clinical deterioration.**

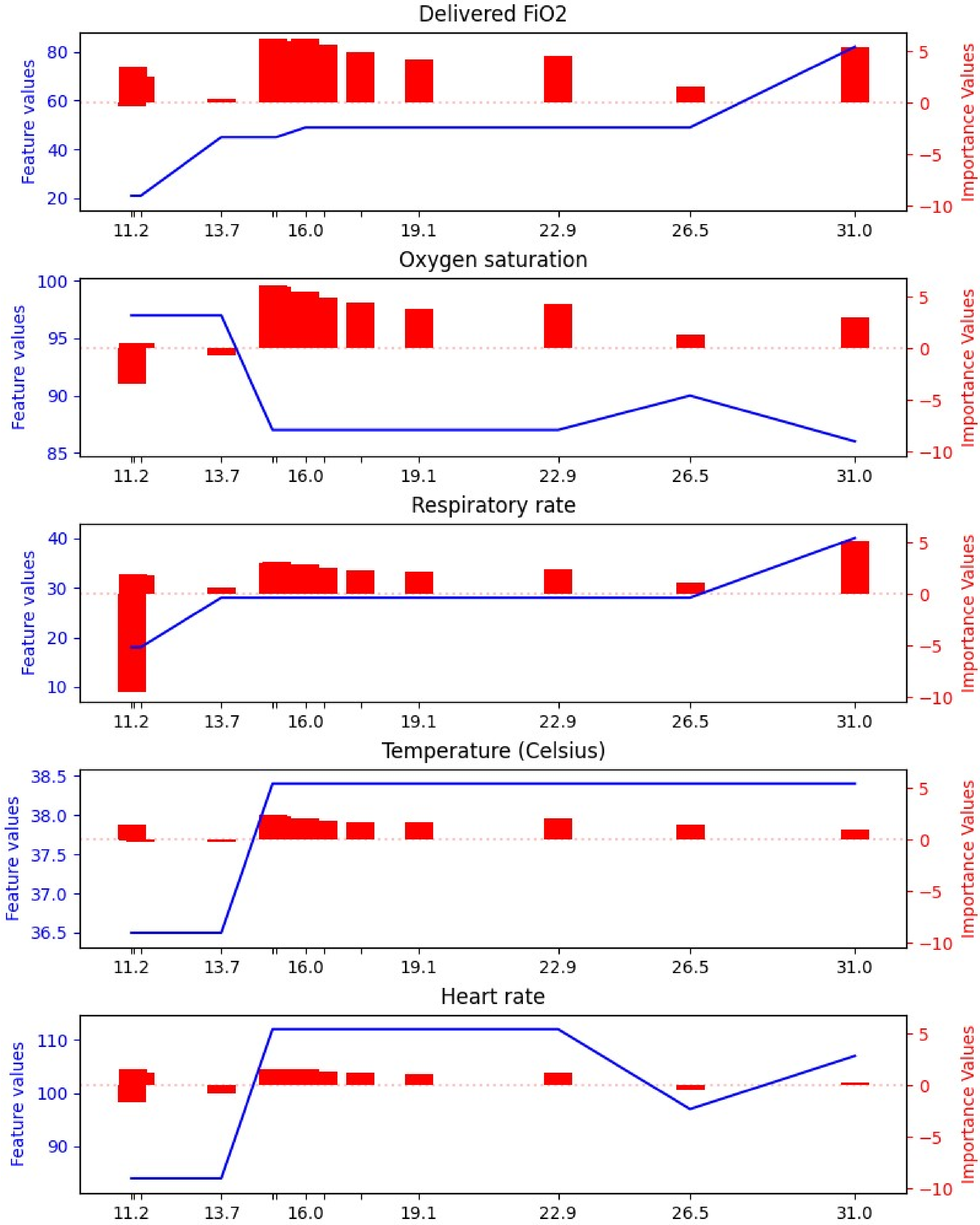
Delivered FiO2
Feature values
Importance Values
80
60
40
20
5
0
−5
−10
11.2
13.7
16.0
19.1
22.9
26.5
31.0
Oxygen saturation
100
95
90
85
Respiratory rate
40
30
20
10
Temperature (Celsius)
38.5
38.0
37.5
37.0
36.5
Heart rate
110
100
90

**Before answering the questions below, would it be helpful to see the vital signs, lab results, and description of the patient again?**

- Yes
- No

**Before answering the questions below, would it be helpful to see the description of an attribution explanation again?**

- Yes
- No

**For the features shown below, please rank the features from the most important (that you would use to make a prediction about this patient's likelihood of clinical deterioration in the next 24 hours) to the least important.**

| (One selection allowed per column) | Most Important (1) | Second-most Important (2) | Third-most Important (3) | Fourth-most Important (4) | Fifth-most Important (5) |
|---|---|---|---|---|---|
| **Heart rate** | ○ | ○ | ○ | ○ | ○ |
| **Respiratory rate** | ○ | ○ | ○ | ○ | ○ |
| **SBP** | ○ | ○ | ○ | ○ | ○ |
| **DBP** | ○ | ○ | ○ | ○ | ○ |
| **Oxygen saturation** | ○ | ○ | ○ | ○ | ○ |
| **Temperature (Celsius)** | ○ | ○ | ○ | ○ | ○ |
| **AVPU mental status** | ○ | ○ | ○ | ○ | ○ |
| **Disoriented** | ○ | ○ | ○ | ○ | ○ |
| **Delivered FiO2** | ○ | ○ | ○ | ○ | ○ |
| **Braden Scale - Nutrition** | ○ | ○ | ○ | ○ | ○ |
| **Sum Total of Braden Scale** | ○ | ○ | ○ | ○ | ○ |
| **Lactate** | ○ | ○ | ○ | ○ | ○ |
| **Creatinine** | ○ | ○ | ○ | ○ | ○ |

**Please select one of the options below to indicate your level of agreement with the statement: "I understand the explanation of the machine learning model's prediction for this patient case."**

| I agree strongly | I agree somewhat | I'm neutral about it | I disagree somewhat | I disagree strongly |
|---|---|---|---|---|
| ○ | ○ | ○ | ○ | ○ |

**Please select one of the options below to indicate your level of agreement with the statement: "The explanation increases my trust in the machine learning model's predictions."**

| I agree strongly | I agree somewhat | I'm neutral about it | I disagree somewhat | I disagree strongly |
|---|---|---|---|---|
| ○ | ○ | ○ | ○ | ○ |

**Please select one of the options below to indicate your level of agreement with the statement: "The explanation increased my understanding of how the machine learning model made a prediction for this patient."**

| | I agree strongly | I agree somewhat | I'm neutral about it | I disagree somewhat | I disagree strongly |
|---|---|---|---|---|---|
| | ○ | ○ | ○ | ○ | ○ |

<< Previous Page

Next Page >>

Save & Return Later

Powered by REDCap

#### User Preferences for Machine Learning Model Explanations

**Page 18 of 26**

### Survey Question 4

**Here we show a counterfactual explanation for why the patient has been identified as being at high risk of clinical deterioration.**

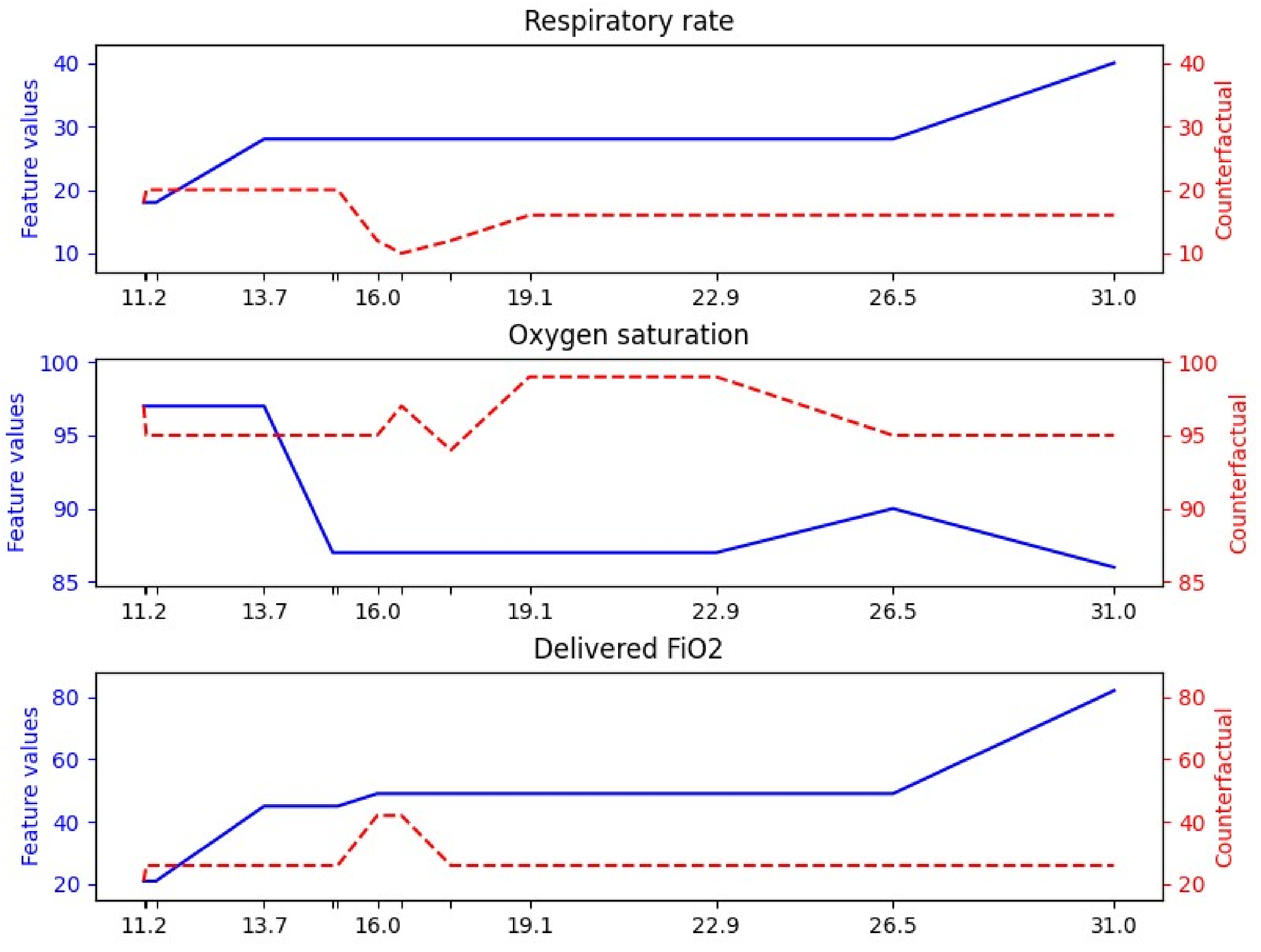


**Before answering the questions below, would it be helpful to see the vital signs, lab results, and description of the patient again?**

- Yes
- No

**Before answering the questions below, would it be helpful to see the description of a counterfactual explanation again?**

- Yes
- No

**For the features shown below, please rank the features from the most important (that you would use to make a prediction about this patient's likelihood of clinical deterioration in the next 24 hours) to the least important.**

| (One selection allowed per column) | Most Important (1) | Second-most Important (2) | Third-most Important (3) | Fourth-most Important (4) | Fifth-most Important (5) |
|---|---|---|---|---|---|
| **Heart rate** | ○ | ○ | ○ | ○ | ○ |
| **Respiratory rate** | ○ | ○ | ○ | ○ | ○ |
| **SBP** | ○ | ○ | ○ | ○ | ○ |
| **DBP** | ○ | ○ | ○ | ○ | ○ |
| **Oxygen saturation** | ○ | ○ | ○ | ○ | ○ |
| **Temperature (Celsius)** | ○ | ○ | ○ | ○ | ○ |
| **AVPU mental status** | ○ | ○ | ○ | ○ | ○ |
| **Disoriented** | ○ | ○ | ○ | ○ | ○ |
| **Delivered FiO2** | ○ | ○ | ○ | ○ | ○ |
| **Braden Scale - Nutrition** | ○ | ○ | ○ | ○ | ○ |
| **Sum Total of Braden Scale** | ○ | ○ | ○ | ○ | ○ |
| **Lactate** | ○ | ○ | ○ | ○ | ○ |
| **Creatinine** | ○ | ○ | ○ | ○ | ○ |

**Please select one of the options below to indicate your level of agreement with the statement: "I understand the explanation of the machine learning model's prediction for this patient case."**

| I agree strongly | I agree somewhat | I'm neutral about it | I disagree somewhat | I disagree strongly |
|---|---|---|---|---|
| ○ | ○ | ○ | ○ | ○ |

**Please select one of the options below to indicate your level of agreement with the statement: "The explanation increases my trust in the machine learning model's predictions."**

| I agree strongly | I agree somewhat | I'm neutral about it | I disagree somewhat | I disagree strongly |
|---|---|---|---|---|
| ○ | ○ | ○ | ○ | ○ |

**Please select one of the options below to indicate your level of agreement with the statement: "The explanation increased my understanding of how the machine learning model made a prediction for this patient."**

| I agree strongly | I agree somewhat | I'm neutral about it | I disagree somewhat | I disagree strongly |
|---|---|---|---|---|
| ○ | ○ | ○ | ○ | ○ |

<< Previous Page

Next Page >>

Save & Return Later

Powered by REDCap

### User Preferences for Machine Learning Model Explanations

A A A

Page 19 of 26

### Survey Question 4

**Here we show a rule-based explanation for why the patient has been identified as being at high risk of clinical deterioration.**

**BECAUSE:**

Respiratory rate at time 31.0 > 20
Delivered FiO2 at time 22.9 > 21
Delivered FiO2 at time 26.5 > 21

**THEN:**

This patient is expected to be at high risk of clinical deterioration.

**Before answering the questions below, would it be helpful to see the vital signs, lab results, and description of the patient again?**

- ○ Yes
- ○ No

**Before answering the questions below, would it be helpful to see the description of a rule-based explanation again?**

- ○ Yes
- ○ No

**For the features shown below, please rank the features from the most important (that you would use to make a prediction about this patient's likelihood of clinical deterioration in the next 24 hours) to the least important.**

| (One selection allowed per column) | Most Important (1) | Second-most Important (2) | Third-most Important (3) | Fourth-most Important (4) | Fifth-most Important (5) |
|---|---|---|---|---|---|
| **Heart rate** | ○ | ○ | ○ | ○ | ○ |
| **Respiratory rate** | ○ | ○ | ○ | ○ | ○ |
| **SBP** | ○ | ○ | ○ | ○ | ○ |

| | | | | | |
|---|---|---|---|---|---|
| **DBP** | ○ | ○ | ○ | ○ | ○ |
| **Oxygen saturation** | ○ | ○ | ○ | ○ | ○ |
| **Temperature (Celsius)** | ○ | ○ | ○ | ○ | ○ |
| **AVPU mental status** | ○ | ○ | ○ | ○ | ○ |
| **Disoriented** | ○ | ○ | ○ | ○ | ○ |
| **Delivered FiO2** | ○ | ○ | ○ | ○ | ○ |
| **Braden Scale - Nutrition** | ○ | ○ | ○ | ○ | ○ |
| **Sum Total of Braden Scale** | ○ | ○ | ○ | ○ | ○ |
| **Lactate** | ○ | ○ | ○ | ○ | ○ |
| **Creatinine** | ○ | ○ | ○ | ○ | ○ |

**Please select one of the options below to indicate your level of agreement with the statement: "I understand the explanation of the machine learning model's prediction for this patient case."**

| I agree strongly | I agree somewhat | I'm neutral about it | I disagree somewhat | I disagree strongly |
|---|---|---|---|---|
| ○ | ○ | ○ | ○ | ○ |

**Please select one of the options below to indicate your level of agreement with the statement: "The explanation increases my trust in the machine learning model's predictions."**

| I agree strongly | I agree somewhat | I'm neutral about it | I disagree somewhat | I disagree strongly |
|---|---|---|---|---|
| ○ | ○ | ○ | ○ | ○ |

**Please select one of the options below to indicate your level of agreement with the statement: "The explanation increased my understanding of how the machine learning model made a prediction for this patient."**

| I agree strongly | I agree somewhat | I'm neutral about it | I disagree somewhat | I disagree strongly |
|---|---|---|---|---|
| ○ | ○ | ○ | ○ | ○ |

<< Previous Page

Next Page >>

Save & Return Later

Powered by REDCap

#### User Preferences for Machine Learning Model Explanations

A A A

**Page 20 of 26**

### Survey Question 5

**You have received an alert that a patient has been identified as being at high risk of clinical deterioration. For this patient, the vital signs, lab results, and a description of the patient are displayed below.**

**Clinical Notes:**

The patient is a 71-year-old male with a BMI of 26.9

**The features given to the machine learning model are shown below.**

| Hours Since Admit | 3.8 | 4.1 | 4.5 | 5.1 | 5.5 | 7 | 8.4 | 19.6 | 22 | 23.8 | 26.[illegible] |
|---|---|---|---|---|---|---|---|---|---|---|---|
| **Vitals** | | | | | | | | | | | |
| **Heart rate** | 88 | - | - | - | - | 113 | 109 | 94 | 108 | - | 13[illegible] |
| **Respiratory rate** | - | 20 | - | - | - | - | 21 | 17 | 21 | 41 | 33 |
| **SBP** | 115 | - | 109 | - | - | 95 | 99 | 116 | - | - | - |
| **DBP** | 73 | - | 72 | - | - | 65 | 69 | 75 | - | - | - |
| **Oxygen saturation** | 97 | - | - | - | - | 94 | 91 | 92 | 92 | - | 74 |
| **Temperature (Celsius)** | 36.8 | - | - | - | - | 36.8 | 36.9 | 37 | 36.2 | - | - |
| **AVPU mental status** | - | - | - | Responds to Pain | - | - | - | - | - | - | - |
| **Disoriented** | - | - | - | No | - | - | - | - | - | - | - |
| **Delivered FiO2** | - | 74 | - | - | - | - | 34 | - | 34 | - | 10[illegible] |
| **Braden Scale - Nutrition** | - | - | - | - | 3 | - | - | - | - | - | - |
| **Sum Total of Braden Scale** | - | - | - | - | 17 | - | - | - | - | - | - |
| **Labs** | | | | | | | | | | | |
| **Lactate** | - | - | - | - | - | - | - | - | - | - | - |
| **Creatinine** | - | - | - | - | - | - | - | - | - | - | - |

**For the features shown, please rank what you think are the five most important features (that you would use to make prediction about this patient's likelihood of clinical deterioration in the next 24 hours).**

| (One selection allowed per column) | Most Important (1) | Second-most Important (2) | Third-most Important (3) | Fourth-most Important (4) | Fifth-most Important ([illegible] |
|---|---|---|---|---|---|
| **Heart rate** | ○ | ○ | ○ | ○ | ○ |
| **Respiratory rate** | ○ | ○ | ○ | ○ | ○ |
| **SBP** | ○ | ○ | ○ | ○ | ○ |
| **DBP** | ○ | ○ | ○ | ○ | ○ |

| | | | | | |
|---|---|---|---|---|---|
| **Oxygen saturation** | ○ | ○ | ○ | ○ | ○ |
| **Temperature (Celsius)** | ○ | ○ | ○ | ○ | ○ |
| **AVPU mental status** | ○ | ○ | ○ | ○ | ○ |
| **Disoriented** | ○ | ○ | ○ | ○ | ○ |
| **Delivered FiO2** | ○ | ○ | ○ | ○ | ○ |
| **Braden Scale - Nutrition** | ○ | ○ | ○ | ○ | ○ |
| **Sum Total of Braden Scale** | ○ | ○ | ○ | ○ | ○ |
| **Lactate** | ○ | ○ | ○ | ○ | ○ |
| **Creatinine** | ○ | ○ | ○ | ○ | ○ |

<< Previous Page

Next Page >>

Save & Return Later

Powered by REDCap

### User Preferences for Machine Learning Model Explanations

A A A



### Survey Question 5

**Here we show a rule-based explanation for why the patient has been identified as being at high risk of clinical deterioration.**

**BECAUSE:**

Respiratory rate at time 26.4 > 20
Delivered FiO2 at time 20.6 > 21
Delivered FiO2 at time 23.8 > 21
Delivered FiO2 at time 26.2 > 21
Delivered FiO2 at time 26.4 > 21

**THEN:**

This patient is expected to be at high risk of clinical deterioration.

**Before answering the questions below, would it be helpful to see the vital signs, lab results, and description of the patient again?**

- Yes
- No

**Before answering the questions below, would it be helpful to see the description of a rule-based explanation again?**

- Yes
- No

**For the features shown below, please rank the features from the most important (that you would use to make a prediction about this patient's likelihood of clinical deterioration in the next 24 hours) to the least important.**

| (One selection allowed per column) | Most Important (1) | Second-most Important (2) | Third-most Important (3) | Fourth-most Important (4) | Fifth-most Important (5) |
|---|---|---|---|---|---|
| **Heart rate** | ○ | ○ | ○ | ○ | ○ |
| **Respiratory rate** | ○ | ○ | ○ | ○ | ○ |

| | | | | | |
|---|---|---|---|---|---|
| **SBP** | ○ | ○ | ○ | ○ | ○ |
| **DBP** | ○ | ○ | ○ | ○ | ○ |
| **Oxygen saturation** | ○ | ○ | ○ | ○ | ○ |
| **Temperature (Celsius)** | ○ | ○ | ○ | ○ | ○ |
| **AVPU mental status** | ○ | ○ | ○ | ○ | ○ |
| **Disoriented** | ○ | ○ | ○ | ○ | ○ |
| **Delivered FiO2** | ○ | ○ | ○ | ○ | ○ |
| **Braden Scale - Nutrition** | ○ | ○ | ○ | ○ | ○ |
| **Sum Total of Braden Scale** | ○ | ○ | ○ | ○ | ○ |
| **Lactate** | ○ | ○ | ○ | ○ | ○ |
| **Creatinine** | ○ | ○ | ○ | ○ | ○ |

**Please select one of the options below to indicate your level of agreement with the statement: "I understand the explanation of the machine learning model's prediction for this patient case."**

| I agree strongly | I agree somewhat | I'm neutral about it | I disagree somewhat | I disagree strongly |
|---|---|---|---|---|
| ○ | ○ | ○ | ○ | ○ |

**Please select one of the options below to indicate your level of agreement with the statement: "The explanation increases my trust in the machine learning model's predictions."**

| I agree strongly | I agree somewhat | I'm neutral about it | I disagree somewhat | I disagree strongly |
|---|---|---|---|---|
| ○ | ○ | ○ | ○ | ○ |

**Please select one of the options below to indicate your level of agreement with the statement: "The explanation increased my understanding of how the machine learning model made a prediction for this patient."**

| I agree strongly | I agree somewhat | I'm neutral about it | I disagree somewhat | I disagree strongly |
|---|---|---|---|---|
| ○ | ○ | ○ | ○ | ○ |

<< Previous Page

Next Page >>

Save & Return Later

Powered by REDCap

#### User Preferences for Machine Learning Model Explanations

A A A

**Page 22 of 26**

### Survey Question 5

**Here we show a counterfactual explanation for why the patient has been identified as being at high risk of clinical deterioration.**

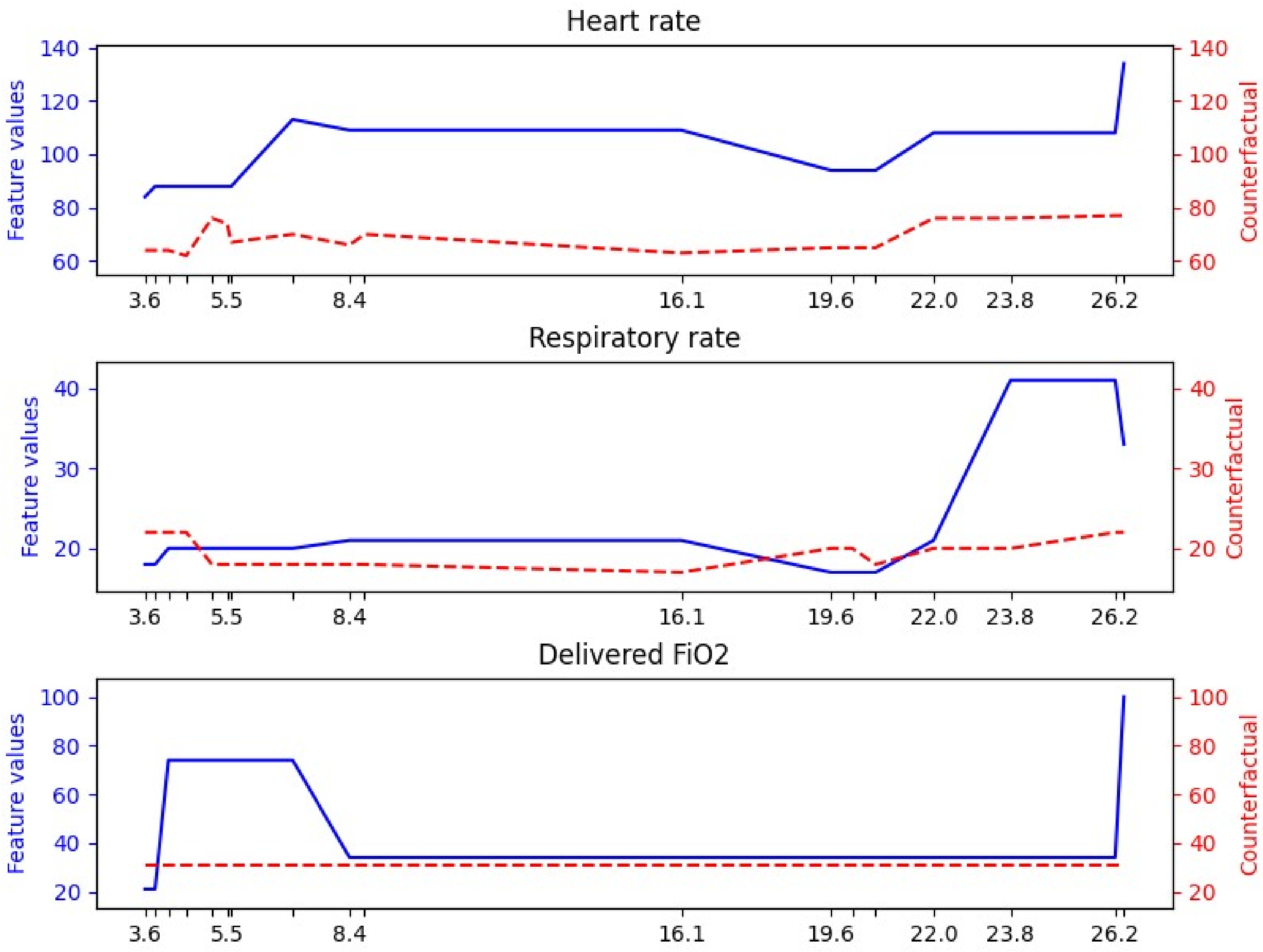


**Before answering the questions below, would it be helpful to see the vital signs, lab results, and description of the patient again?**

- Yes
- No

**Before answering the questions below, would it be helpful to see the description of a counterfactual explanation again?**

- Yes
- No

**For the features shown below, please rank the features from the most important (that you would use to make a prediction about this patient's likelihood of clinical deterioration in the next 24 hours) to the least important.**

| (One selection allowed per column) | Most Important (1) | Second-most Important (2) | Third-most Important (3) | Fourth-most Important (4) | Fifth-most Important (5) |
|---|---|---|---|---|---|
| **Heart rate** | ○ | ○ | ○ | ○ | ○ |
| **Respiratory rate** | ○ | ○ | ○ | ○ | ○ |
| **SBP** | ○ | ○ | ○ | ○ | ○ |
| **DBP** | ○ | ○ | ○ | ○ | ○ |
| **Oxygen saturation** | ○ | ○ | ○ | ○ | ○ |
| **Temperature (Celsius)** | ○ | ○ | ○ | ○ | ○ |
| **AVPU mental status** | ○ | ○ | ○ | ○ | ○ |
| **Disoriented** | ○ | ○ | ○ | ○ | ○ |
| **Delivered FiO2** | ○ | ○ | ○ | ○ | ○ |
| **Braden Scale - Nutrition** | ○ | ○ | ○ | ○ | ○ |
| **Sum Total of Braden Scale** | ○ | ○ | ○ | ○ | ○ |
| **Lactate** | ○ | ○ | ○ | ○ | ○ |
| **Creatinine** | ○ | ○ | ○ | ○ | ○ |

**Please select one of the options below to indicate your level of agreement with the statement: "I understand the explanation of the machine learning model's prediction for this patient case."**

| I agree strongly | I agree somewhat | I'm neutral about it | I disagree somewhat | I disagree strongly |
|---|---|---|---|---|
| ○ | ○ | ○ | ○ | ○ |

**Please select one of the options below to indicate your level of agreement with the statement: "The explanation increases my trust in the machine learning model's predictions."**

| I agree strongly | I agree somewhat | I'm neutral about it | I disagree somewhat | I disagree strongly |
|---|---|---|---|---|
| ○ | ○ | ○ | ○ | ○ |

**Please select one of the options below to indicate your level of agreement with the statement: "The explanation increased my understanding of how the machine learning model made a prediction for this patient."**

| I agree strongly | I agree somewhat | I'm neutral about it | I disagree somewhat | I disagree strongly |
|---|---|---|---|---|
| ○ | ○ | ○ | ○ | ○ |

<< Previous Page

Next Page >>

Save & Return Later

Powered by REDCap

#### User Preferences for Machine Learning Model Explanations

AAA

Page 23 of 26

### Survey Question 5

**Here we show an attribution explanation for why the patient has been identified as being at high risk of clinical deterioration.**

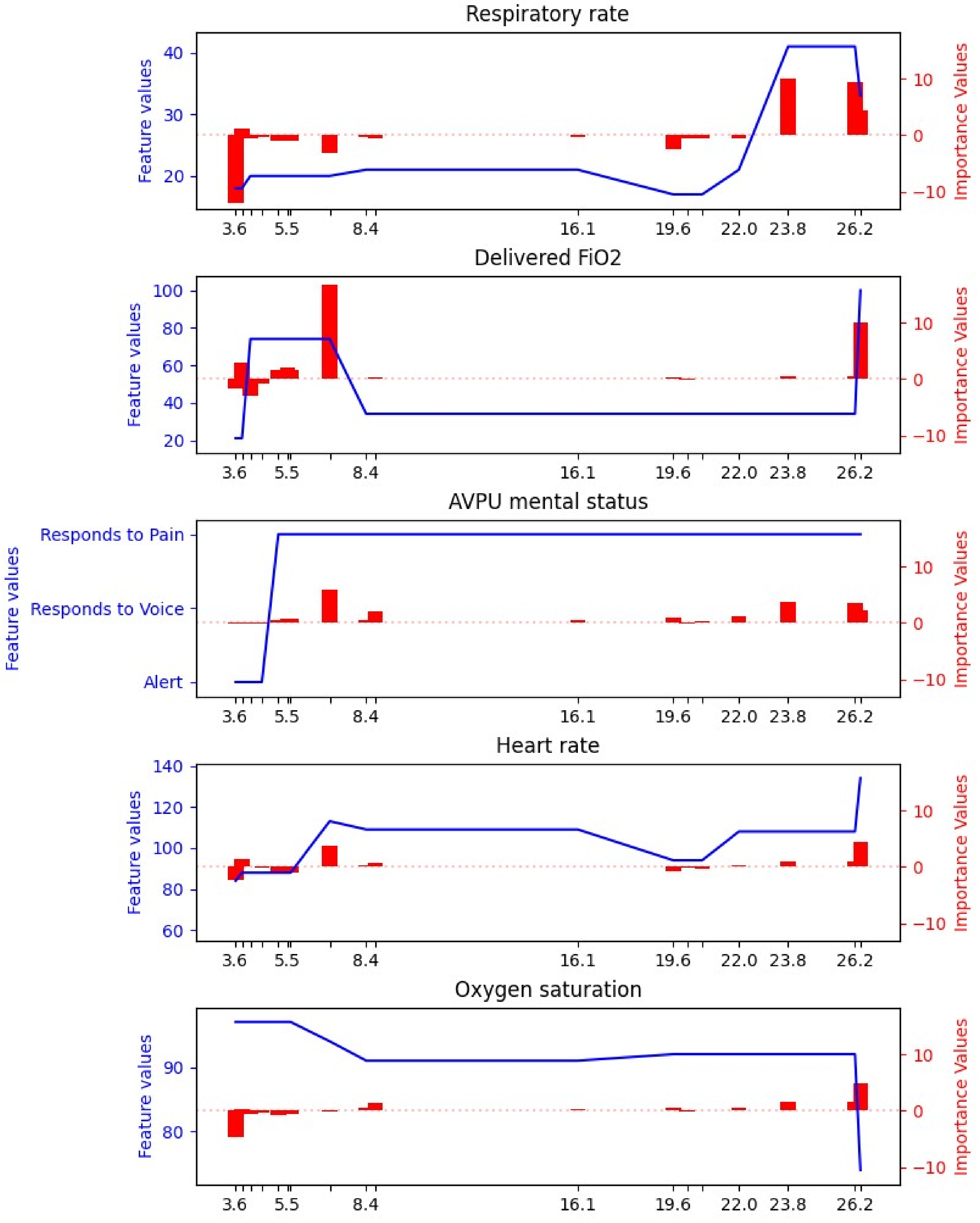
Respiratory rate
Feature values
Importance Values
Delivered FiO2
AVPU mental status
Responds to Pain
Responds to Voice
Alert
Heart rate
Oxygen saturation
3.6
5.5
8.4
16.1
19.6
22.0
23.8
26.2

**Before answering the questions below, would it be helpful to see the vital signs, lab results, and description of the patient again?**

- ○ Yes
- ○ No

**Before answering the questions below, would it be helpful to see the description of an attribution explanation again?**

- ○ Yes
- ○ No

**For the features shown below, please rank the features from the most important (that you would use to make a prediction about this patient's likelihood of clinical deterioration in the next 24 hours) to the least important.**

| (One selection allowed per column) | Most Important (1) | Second-most Important (2) | Third-most Important (3) | Fourth-most Important (4) | Fifth-most Important (5) |
|---|---|---|---|---|---|
| **Heart rate** | ○ | ○ | ○ | ○ | ○ |
| **Respiratory rate** | ○ | ○ | ○ | ○ | ○ |
| **SBP** | ○ | ○ | ○ | ○ | ○ |
| **DBP** | ○ | ○ | ○ | ○ | ○ |
| **Oxygen saturation** | ○ | ○ | ○ | ○ | ○ |
| **Temperature (Celsius)** | ○ | ○ | ○ | ○ | ○ |
| **AVPU mental status** | ○ | ○ | ○ | ○ | ○ |
| **Disoriented** | ○ | ○ | ○ | ○ | ○ |
| **Delivered FiO2** | ○ | ○ | ○ | ○ | ○ |
| **Braden Scale - Nutrition** | ○ | ○ | ○ | ○ | ○ |
| **Sum Total of Braden Scale** | ○ | ○ | ○ | ○ | ○ |
| **Lactate** | ○ | ○ | ○ | ○ | ○ |
| **Creatinine** | ○ | ○ | ○ | ○ | ○ |

**Please select one of the options below to indicate your level of agreement with the statement: "I understand the explanation of the machine learning model's prediction for this patient case."**

| I agree strongly | I agree somewhat | I'm neutral about it | I disagree somewhat | I disagree strongly |
|---|---|---|---|---|
| ○ | ○ | ○ | ○ | ○ |

**Please select one of the options below to indicate your level of agreement with the statement: "The explanation increases my trust in the machine learning model's predictions."**

| I agree strongly | I agree somewhat | I'm neutral about it | I disagree somewhat | I disagree strongly |
|---|---|---|---|---|
| ○ | ○ | ○ | ○ | ○ |

**Please select one of the options below to indicate your level of agreement with the statement: "The explanation increased my understanding of how the machine learning model made a prediction for this patient."**

| | I agree strongly | I agree somewhat | I'm neutral about it | I disagree somewhat | I disagree strongly |
|---|---|---|---|---|---|
| | ○ | ○ | ○ | ○ | ○ |

<< Previous Page

Next Page >>

Save & Return Later

Powered by REDCap

### User Preferences for Machine Learning Model Explanations

AAA



### Concluding Questions

**For each explanation type listed on the left, please select one of the options below to indicate your level of agreement with the statement: "I understand explanations of this type."**

| | I agree strongly | I agree somewhat | I'm neutral about it | I disagree somewhat | I disagree strongly |
|---|---|---|---|---|---|
| Attribution explanations | ○ | ○ | ○ | ○ | ○ |
| Counterfactual explanations | ○ | ○ | ○ | ○ | ○ |
| Rule-based explanations | ○ | ○ | ○ | ○ | ○ |

**For each explanation type listed on the left, please select one of the options below to indicate your level of agreement with the statement: "Explanations of this type increased my trust in the machine learning model."**

| | I agree strongly | I agree somewhat | I'm neutral about it | I disagree somewhat | I disagree strongly |
|---|---|---|---|---|---|
| Attribution explanations | ○ | ○ | ○ | ○ | ○ |
| Counterfactual explanations | ○ | ○ | ○ | ○ | ○ |
| Rule-based explanations | ○ | ○ | ○ | ○ | ○ |

**For each explanation type listed on the left, please select one of the options below to indicate your level of agreement with the statement: "Explanations of this type increased my understanding of how the machine learning model makes predictions for patients."**

| | I agree strongly | I agree somewhat | I'm neutral about it | I disagree somewhat | I disagree strongly |
|---|---|---|---|---|---|
| Attribution explanations | ○ | ○ | ○ | ○ | ○ |
| Counterfactual explanations | ○ | ○ | ○ | ○ | ○ |
| Rule-based explanations | ○ | ○ | ○ | ○ | ○ |

**For each explanation type listed on the left, please select one of the options below to indicate your level of agreement with the statement: "Explanations of this type influenced my selection of which features were most important for determining that a patient was at high risk of clinical**

**deterioration."**

| | I agree strongly | I agree somewhat | I'm neutral about it | I disagree somewhat | I disagree strongly |
|---|---|---|---|---|---|
| **Attribution explanations** | ○ | ○ | ○ | ○ | ○ |
| **Counterfactual explanations** | ○ | ○ | ○ | ○ | ○ |
| **Rule-based explanations** | ○ | ○ | ○ | ○ | ○ |

**Please select one of the options below to indicate your level of agreement with the statement: "It is important to see explanations of predictions for machine learning models that are deployed in clinical settings."**

| I agree strongly | I agree somewhat | I'm neutral about it | I disagree somewhat | I disagree strongly |
|---|---|---|---|---|
| ○ | ○ | ○ | ○ | ○ |

**Would you prefer to see multiple types of explanations together for a given patient?**

| Yes, I would like to see multiple explanation types together | No, I want to see only one explanation type at a time |
|---|---|
| ○ | ○ |

**Please select one of the options below to indicate your level of agreement with the statement: "For rule-based explanations, I would prefer to see a shorter rule with fewer conditions that is less representative of the model over a more complex over a more complex rule with more conditions that is closer to the true representation of the model."**

| I agree strongly | I agree somewhat | I'm neutral about it | I disagree somewhat | I disagree strongly |
|---|---|---|---|---|
| ○ | ○ | ○ | ○ | ○ |

**Please select one of the options below to indicate your level of agreement with the statement: "For an attribution explanation, I would prefer to see only the top few most important features and their importance values instead of all of the features and their importance values."**

| I agree strongly | I agree somewhat | I'm neutral about it | I disagree somewhat | I disagree strongly |
|---|---|---|---|---|
| ○ | ○ | ○ | ○ | ○ |

What do you like or dislike about attribution explanations?

What do you like or dislike about counterfactual explanations?

What do you like or dislike about rule-based explanations?

<< Previous Page

Next Page >>

Save & Return Later

Powered by REDCap

### User Preferences for Machine Learning Model Explanations

A A A

Page 25 of 26

#### Information

**Which of the two institutions below are you affiliated with?**
* must provide value

- UW-Madison
- UChicago

**What is your gender?**

- Female
- Male
- Another gender not listed above

**What is your age range?**

- 20-29 years old
- 30-39 years old
- 40-49 years old
- 50-59 years old
- 60-years or older

**How would you describe your race or ethnicity (choose all that apply)?**

- Alaska Native
- Asian
- Black or African American
- Hispanic or Latinx
- Native American or American Indian
- Native Hawaiian or Pacific Islander
- White
- Another race or ethnicity not listed above

**Which of the following describes your current role?**

- Nurse
- Rapid response team member
- Physician Trainee
- Attending physician
- Mid-level provider

**How many years of clinical experience do you have?**

- Less than 1 year
- 1-5 years
- >5-10 years
- >10 years

**These last three questions are required in order to reimburse you for your time.**

**What is your Full Name?**

* must provide value

**What is your Email Address?**

* must provide value

<< Previous Page

Next Page >>

Save & Return Later

Powered by REDCap

### User Preferences for Machine Learning Model Explanations

A A A

**Page 26 of 26**

#### General Comments

**Do you have any general comments?**

**In a few business days, you will receive an email to set up an account and enter all of the bank information needed for the ACH payment.**

**Thank you for your time!**

<< Previous Page

Submit

Save & Return Later

Powered by REDCap
